# Current status of methodological and reporting quality of systematic reviews and meta-analyses in medicine and health science fields in Ethiopia: Leveraging quantity to improve quality

**DOI:** 10.1101/2022.05.16.22275144

**Authors:** Tesfa Dejenie Habtewold, Nigussie Tadesse Sharew, Aklilu Endalamaw, Henok Mulugeta, Getenet Dessie, Getachew Mullu Kassa, Wubet Alebachew Bayih, Mulugeta Molla Birhanu, Andreas A. Teferra, Balewgize Sileshi Tegegne, Nigus G. Asefa, Abera Kenay Tura, Sisay Mulugeta Alemu

**Author notes:** Corresponding author Tesfa Dejenie Habtewold, PhD, Branch of Epidemiology, Division of Population Health Research, Division of Intramural Research, Eunice Kennedy Shriver National Institute of Child Health and Human Development, National Institutes of Health, Bethesda, Maryland, United States of America.

## Abstract

**Introduction:** Despite the rise in the number of systematic reviews (SR) and meta-analyses (MA) in medicine and health science fields in Ethiopia, there is limited up-to-date evidence on their methodological and reporting quality for using them in decision-making. The aim of this study was to characterize epidemiological trends and evaluate the methodological and reporting quality of SR and MA in Ethiopia.

**Methods:** A retrospective observational overview study was conducted on SR and MA in medicine and health science fields in Ethiopia that were accessed through PubMed, PsycINFO, EMBASE, CINAHL databases and additional manual searching. SR and MA based on primary human studies associated with the Ethiopian population, irrespective of the place of publication and authors’ affiliation, and published until March 16, 2021, were included. Title/abstract and full-text screening were conducted in duplicate using EndNote and Covidence semi-automated reference management tools. Data extraction tool was developed using Preferred Reporting Items for Systematic Reviews and Meta-Analyses (PRISMA) and second version of A Measurement Tool to Assess Systematic Reviews (AMSTAR-2) guides. We summarized the data using frequencies and median. Two-tailed Chi-Square and Fisher’s Exact tests for categorical variables were used while Kruskal-Wallis test for quantitative variables at alpha level 0.05 to compare the differences in the background characteristics of SR and MA as well as across the publication years. All analyses were done using R version 4.0.2 for macOS.

**Results:** Of the total 3,125 records initially identified, 349 articles were included in our analyses. Of these, 48 (13.75%) were SR and 301 (86.25%) were MA. The publication rate was dramatically increased with nearly three-quarters (73.9%) of SR and MA published after 2018. Most of the SR and MA included observational studies (92.8%), and infectious disease was the most researched (20.9%) subject area. Number of authors, number of affiliations, publication year, protocol registration, number of primary studies, number of references, citation counts and journal quality were significantly different between SR and MA (p < 0.05). Both SR and MA had a low methodological and reporting quality even though there were improvements in registering protocols, searching databases, and transparently reporting search strategy.

**Conclusions:** The production of SR and MA in Ethiopia has been increased over time, especially during the last three years. There is a promising trend of improvement in methodological and reporting quality even though there is much more to do. This study provides an up-to-date overview of the landscape of SR and MA publication rate and quality leverage in Ethiopia. Authors should equally prioritize quality in addition to the fast-track publication.

## Introduction

With the rise in the number of productions of evidence, the need for summarized synthesis of evidence for healthcare decision-making is becoming evident. Systematic reviews (SR) and meta-analyses (MA) are essential tools to qualitatively and quantitatively synthesize a broad range of evidence for researchers and clinicians that can be accessed within a short time.^1^ Thus, they are indispensable for evidence-based medicine and medical decision-making in clinical practice. As a result, SR and MA became increasingly popular to synthesize evidence from primary studies and the number of SR and MA being published has increased steadily over recent years.^2^ For example, 28,959 articles were tagged as SR in MEDLINE in 2014^3^ and 167,029 articles were indexed as SR in 2021.

With the growing publication rate of SR and MA, quality is a main concern; as a result, an overview of SR and MA can be helpful to characterize and monitor these publications, summarize the main findings and evaluate their methodological and reporting qualities.^2-7^ Overview of SR and MA is a review of previously published SR and/or MA, representing the highest levels of evidence synthesis especially when it is based on randomized controlled trial studies.^2^ An overview studies can be an overview of reviews of interventions and associations, diagnostic test accuracy, economic evaluation, and overviews of systematic reviews of qualitative studies.^8^

To date, four overview studies were conducted in Ethiopia. The first overview study, which examined the trends and methodological quality of 35 SR and 17 MA published until 2018, showed that three-fourths of the studies had poor methodological quality.^9^ The second and third overview studies were conducted on SR and MA of child nutrition (nine articles included)^10^ and birth asphyxia (four articles included)^11^ found moderate methodological quality. Recently, the fourth overview study was conducted by our group using 422 SR and MA that were published before December 31, 2021, and provided a detailed bibliometric information on the publication landscape of SR and MA in Ethiopia.^12^ However, methodological and reporting quality of these SR and MA were not assessed.

Given the large body of SR and MA articles being generated in Ethiopia and their essential role in decision-making, a detailed analysis of epidemiological trends, and methodological and reporting characteristics would meaningfully inform decision making and priority setting. In addition, detailed assessments of background information, and methodological and reporting quality of SR and MA was not done in previously published overviews.^9-12^ Thus, this study aimed to assess the epidemiological trends, and methodological and reporting quality SR and MA in medicine and health science fields in Ethiopia to show the landscape of SR and MA studies and enhance the quality of SR and MA in Ethiopia and other low- and middle-income countries.

## Methods and Materials

### Protocol and registration

The planned search strategy, inclusion and exclusion criteria, study screening and selelction, and statistical methods for data analyses were decided prior to the study. The full project package and the protocol was registered in the Open Science Framework (OSF) (<u>10.17605/osf.io/vapzx</u>). All the supplementary methods and data are available in the publically open database in OSF (https://osf.io/q5dw2).

### Search strategy

A retrospective observational overview study was conducted using SR and MA associated with Ethiopia irrespective of place of publication and authors’ affiliation. We searched SR and MA indexed in PubMed (NCBI), PsycInfo (EBSCOhost), CINAHL (EBSCOhost) and EMBASE (direct access) databases from inception to March 16, 2021. We searched “Ethiopia” and “Ethiop*” terms combined with “OR’ Boolean operator in the title, abstract and keywords of records in PubMed, PsycINFO, EMBASE, and CINAHL. Then, the search was further filtered by article type (i.e., meta-analysis, review, systematic review) and species (i.e., human) in PubMed, methodology (i.e., metasynthesis, meta analysis, systematic review, literature review) in PsycINFO, study type (i.e., systematic review, meta-analysis, human) in EMBASE and publication type (i.e., meta-analysis, meta synthesis, review, systematic review) in CINAHL. To retrieve potentially relevant missing studies, our database search was supplemented by hand searching of tables of content of local journals, such as Ethiopian Medical journal and Ethiopian Journal of Reproductive Health. Grey literature and unpublished/preprint SR and MA databases were not searched given that the quality can be different before and after full peer review.

### Inclusion and exclusion criteria

SR and MA that fulfilled the following criteria were included. First, the article title must be identified as a SR and MA by the author(s). For articles that could not be identified as SR and MA or ambiguous, we inspected relevant information in the methods and results section, and consulted Cochrane ^13^ and PRISMA^14^ guidelines to decide the inclusion of the article. Second, SR and MA must be based on primary studies in medicine and health science fields among Ethiopian population irrespective of the place of publication and authors’ affiliation. Original and updated versions, and duplicates (i.e., only the title or topic is similar) of SR and MA were considered as separate publications and were included in our analysis as they have different publication dates, separate number of citations, and include different authors and affiliations. SR and MA protocols, non-systematic reviews (e.g., scoping, historic, literature, or narrative reviews), exact duplicates (i.e., all the title and authors are the same), conference abstracts, grey literature, commentaries and letters to the editors, reviews following case reports, and SR and MA in non-human research subjects were excluded. In addition, SR and MA based on non-medical and -health science topics, and international primary studies were excluded. Furthermore, SR and MA without full text were excluded after contacting corresponding authors, searching in ResearchGate, or searching in free scientific article downloading sites. Articles were classified as MA when SR combined with quantitative synthesis of estimates, otherwise, they we classified as SR.

### Screening, selection, and data extraction

All retrieved records from the database searches were imported to EndNote X9 software^15^ and then to Covidence, a web-based systematic review management system^16^ for removing duplicates and screening and selection. First, duplicates were automatically removed by Covidence and when not detected, manual removal of duplicates was also done by the authors. Then, double-blinded title and abstract screening was done by two independent reviewers (TD and SM) using Covidence. Next, each SR and MA full-text file was downloaded using EndNote X9 software^15^ and imported to Covidence again. Afterwards, a double-blinded full-text review was also done by two independent reviewers (TD and NT) based on prior-specified inclusion and exclusion criteria. The priority of each criterion to exclude articles is presented as follows: full-text not accessible, non-systematic reviews, continental or worldwide SR and MA, and non-human study subjects. Disagreements during these steps were resolved by discussion and involvement of a third reviewer when necessary. Background and methodological data were extracted using the Google form created by the review authors (TD, NT, SM, AE, HM, GD, GM, NGA, WA). Ten percent of the extracted data were validated. Details on the list of variables and their description was presented in Supplementary Table 1. The data extraction form was developed based on PRIMA 2020 reporting guideline^14^, AMSTAR-2 tool^17^, Cochrane guideline^13^, JBI manual^18^, previous similar studies^19, 20^ and authors expertise. The data extraction form was developed, and new variables were included whenever the information is relevant. For example, citation count, impact factor without self-citation, number of reference and institutional rank were added during the data collection.

### Data analysis

We summarized background characteristics, and methodological and reporting qualities using frequencies with percentages and median with range. Two-tailed Chi-Square and Fisher’s Exact tests for categorical variables and Kruskal-Walis test for quantitative variables at alpha level 0.05 were used to compare the difference in background characteristics between SR and MA as well as across the publication years over time. Fisher’s Exact tests for categorical variables when the assumptions of Chi-Square were not fulfilled. Kruskal-Wallis test was used because of that all the quantitative variables were not normally distributed. All the analyses were done using R version 4.0.2 for macOS.

## Results

### Search results

In total, 3,087 records were retrieved through searching PubMed (n = 1,357), EMBASE (n = 1,174), CINAHL (n = 439) and PsycINFO (n = 117) databases. After removing duplicates (n = 842), 2,245 titles and abstracts were screened and 405 were selected for full-text review. Exclusion reasons included regional or international systematic reviews SR and MA, non-related titles, case reports, primary studies, protocols, non-systematic reviews, and commentaries, corrections, and editorials. In additional, five SR and MA were excluded since full-texts were not retrieved after several attempts. After full-text review, 63 regional or international, 10 non-systematic, and three animal study SR and MA were excluded. Through hand searching of the table of contents of local journals and Google, we found an additional 38 records and 25 of them fulfilled our inclusion criteria. Finally, 349 SR and MA were included in the final analysis. The PRISMA flow diagram of the screening and selection process of the identified studies is shown in Fig. 1.

**Fig. 1:**
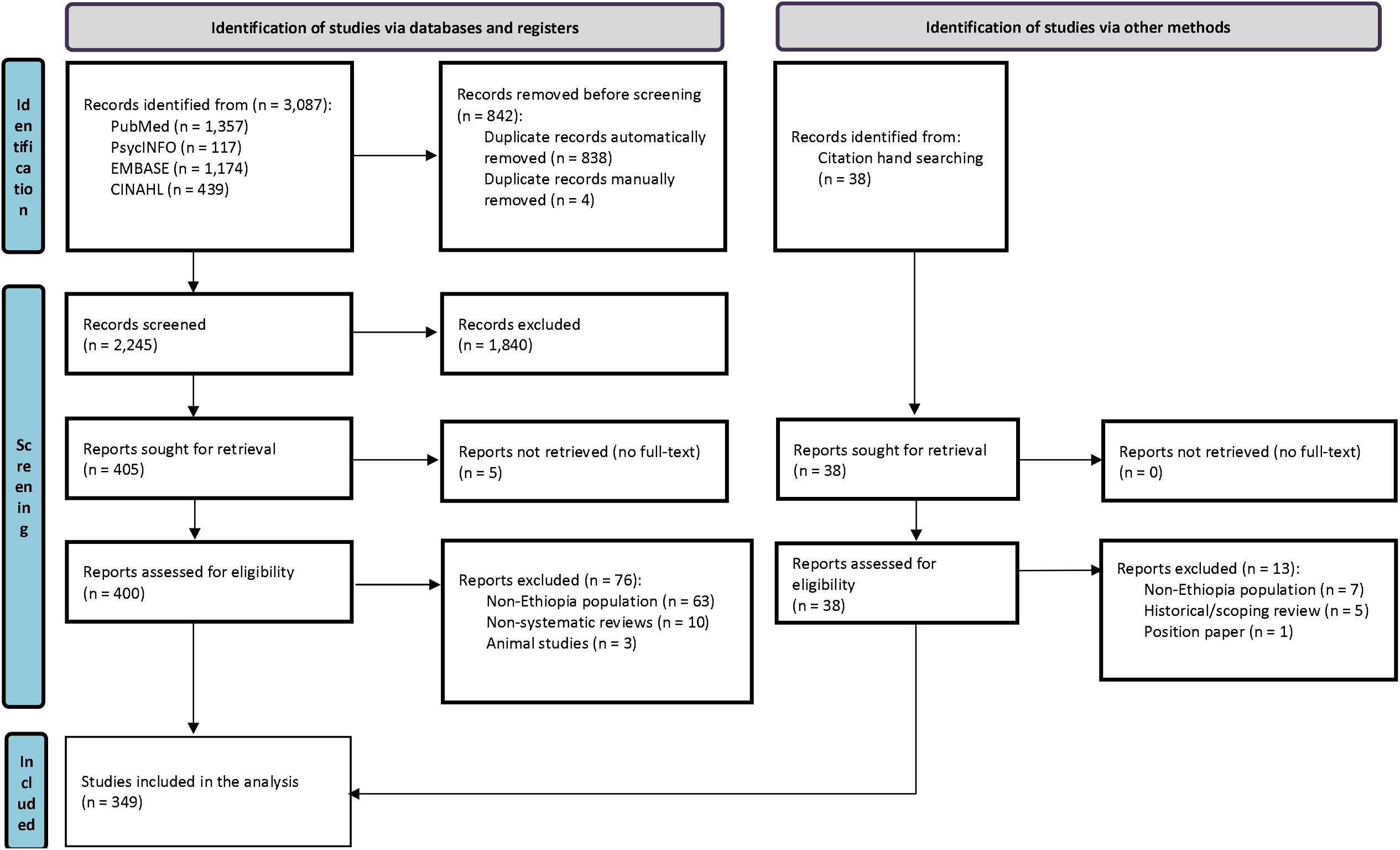
PRISMA flow diagram of literature identification, screening, and selection process.

### Epidemiological trends and characteristics of SR and MA

In total, 48 (13.75%) SR and 301 (86.25%) MA were published with dramatic increment of annual publication rates. Nearly three-quarters of SR and MA (73.9%) were published since 2018 with the highest number of SR and MA (57.0%) published by authors affiliated to institutions in Amhara Region, Ethiopia. University of Gondar (18.9%) is the leading university (Fig 2.) and Nursing department (24.9%) is the most active department in publishing SR and MA (Fig. 3). Infectious diseases (20.9%) were the most published subject areas (Fig. 4). The median number of authors, citation counts, primary studies and cited references was 4, 6, 17 and 54 respectively. The highest number of studies (37.5%) were published in Q2 (25 to 50% group) journals while more than a quarter (26.9%) of SR and MA were published in unranked journals. BioMed Central (43.8%) and PLOS ONE (10.3%) were the most chosen publisher and journal, respectively (Table 1).

**Table 1:** Background information of SR and MA (N=349).

| Characteristics | Article type |  | Total (N=349) | W/X<sup>2</sup> | p-value |
| --- | --- | --- | --- | --- | --- |
|  | Systematic reviews (N=48) | Meta-analyses (N=301) |  |  |  |
| **Number of authors, median (range)** | 3.00 (12.0) | 4.00 (15.0) | 4.00 (15.0) | 4,530.5 | **<0.0001** |
| **Number of affiliations** |  |  |  |  |  |
| Multiple (>1) | 11 (22.9%) | 29 (9.6%) | 40 (11.5%) | 7.197 | **0.007** |
| Single | 37 (77.1%) | 272 (90.4%) | 309 (88.5%) |  |  |
| **Rank of universities, median (range)** | 5.00 (36.0) | 9.00 (38.0) | 8.00 (38.0) | 4,189.0 | 0.165 |
| **Top 5 most active regions** |  |  |  |  |  |
| Addis Ababa | 8 (16.7%) | 23 (7.6%) | 31 (8.9%) |  |  |
| Amhara | 14 (29.2%) | 185 (61.5%) | 199 (57.0%) |  |  |
| Oromia | 6 (12.5%) | 28 (9.3%) | 34 (9.7%) |  |  |
| SNNPR | 5 (10.4%) | 21 (7.0%) | 26 (7.4%) |  |  |
| Tigray | 4 (8.3%) | 11 (3.7%) | 15 (4.3%) |  |  |
| **Country of affiliation** |  |  |  |  |  |
| Ethiopia | 40 (83.3%) | 286 (95.0%) | 326 (93.4%) | 0.264 | **0.007** |
| International | 8 (16.7%) | 15 (5.0%) | 23 (6.6%) |  |  |
| **Publication year** |  |  |  |  |  |
| ≤2018 | 22 (45.8%) | 69 (22.9%) | 91 (26.1%) | 23.994 | **<0.0001** |
| 2019 | 16 (33.3%) | 56 (18.6%) | 72 (20.6%) |  |  |
| 2020 | 9 (18.8%) | 139 (46.2%) | 148 (42.4%) |  |  |
| 2021 | 1 (2.1%) | 37 (12.3%) | 38 (10.9%) |  |  |
| **Top 5 most active journals** |  |  |  |  |  |
| BMC Infectious Diseases | 1 (2.1%) | 25 (8.3%) | 26 (7.4%) |  |  |
| BMC Public Health | 1 (2.1%) | 16 (5.3%) | 17 (4.9%) |  |  |
| Reproductive Health | 1 (2.1%) | 10 (3.3%) | 11 (3.2%) |  |  |
| BMC Pregnancy and Childbirth | 0 (0%) | 12 (4.0%) | 12 (3.4%) |  |  |
| PLOS ONE | 0 (0%) | 36 (12.0%) | 36 (10.3%) |  |  |
| **Top 6 most active publishers** |  |  |  |  |  |
| BioMed Central | 11 (22.9%) | 142 (47.2%) | 153 (43.8%) |  |  |
| Elsevier | 5 (10.4%) | 20 (6.6%) | 25 (7.2%) |  |  |
| Hindawi | 7 (14.6%) | 28 (9.3%) | 35 (10.0%) |  |  |
| Springer | 2 (4.2%) | 16 (5.3%) | 18 (5.2%) |  |  |
| PLOS | 0 (0%) | 37 (12.3%) | 37 (10.6%) |  |  |
| SAGE | 0 (0%) | 11 (3.7%) | 11 (3.2%) |  |  |
| **2-IF, median (range)** | 1.36 (4.89) | 2.88 (10.6) | 2.69 (10.6) | 5,488.5 | **0.009** |
| **2-IF without self-citation, median (range)** | 1.27 (4.65) | 2.72 (10.2) | 2.52 (10.2) | 5,321.0 | **0.004** |
| **5-year impact factor, median (range)** | 1.46 (5.17) | 3.40 (10.4) | 3.26 (10.4) | 5,258.0 | **0.003** |
| **Journals rank** |  |  |  |  |  |
| Q1 (>75% journal group) | 3 (6.3%) | 25 (8.3%) | 28 (8.0%) | 23.994 | **<0.0001** |
| Q2 (50-75% journal group) | 12 (25.0%) | 119 (39.5%) | 131 (37.5%) |  |  |
| Q3 (25-50% journal group) | 8 (16.7%) | 68 (22.6%) | 76 (21.8%) |  |  |
| Q4 (<25% journal group) | 4 (8.3%) | 13 (4.3%) | 17 (4.9%) |  |  |
| Not ranked | 21 (43.8%) | 73 (24.3%) | 94 (26.9%) |  |  |
| **Protocol registration** |  |  |  |  |  |
| No | 46 (95.8%) | 203 (67.4%) | 249 (71.3%) | 16.323 | **<0.0001** |
| Yes | 2 (4.2%) | 98 (32.6%) | 100 (28.7%) |  |  |
| **Number of primary studies, median (range)** | 22.5 (227.0) | 16.0 (227.0) | 17.0 (227.0) | 8,706.5 | **0.003** |
| **Type of studies included** |  |  |  |  |  |
| All type of studies | 1 (2.1%) | 8 (2.6%) | 9 (2.6%) |  |  |
| Both observational and RCT studies | 1 (2.1%) | 4 (1.3%) | 5 (1.4%) |  |  |
| Observational studies | 39 (81.3%) | 285 (94.7%) | 324 (92.8%) |  |  |
| **Number of references, median (range)** | 46.0 (130.0) | 55.0 (128.0) | 54.0 (128.0) | 5,574.0 | **0.011** |
| **Citation count, median (range)** | 8.50 (193.0) | 6.00 (93.0) | 6.00 (193) | 8,505.5 | **0.048** |
| **Citation count without self-citation, median (range)** | 8.50 (190.0) | 5.00 (93.0) | 6.00 (190.0) | 8,586.0 | **0.035** |
| **Citation manager software\*** |  |  |  |  |  |
| EndNote | 7 (14.6%) | 149 (49.5%) | 156 (44.7%) |  |  |
| Mendeley | 3 (6.3%) | 4 (1.3%) | 7 (2.0%) |  |  |
| RefWorks | 1 (2.1%) | 4 (1.3%) | 5 (1.4%) |  |  |
| Not reported | 37 (77.1%) | 144 (47.8%) | 181 (51.9%) |  |  |

W = Wilcoxon test statistic; $\chi^2$ = Chi-Square test statistic; RCT = Randomized controlled trials; SNNPR = Southern Nations, Nationalities, and People's Region; BMC = BioMed Central; HIV/AIDS = Human immunodeficiency virus infection and acquired immunodeficiency syndrome

**Fig 2.**
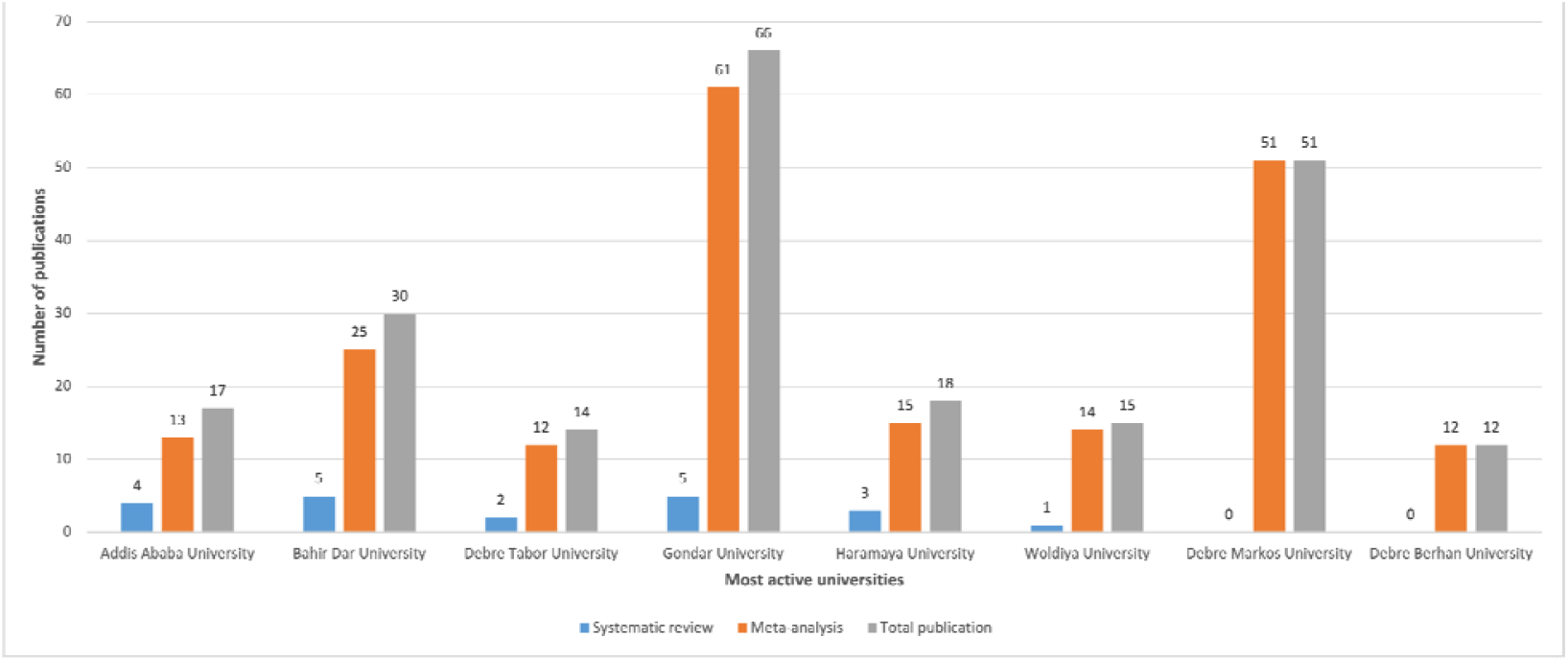
Most active universities publishing SR and MA.

**Fig 3.**
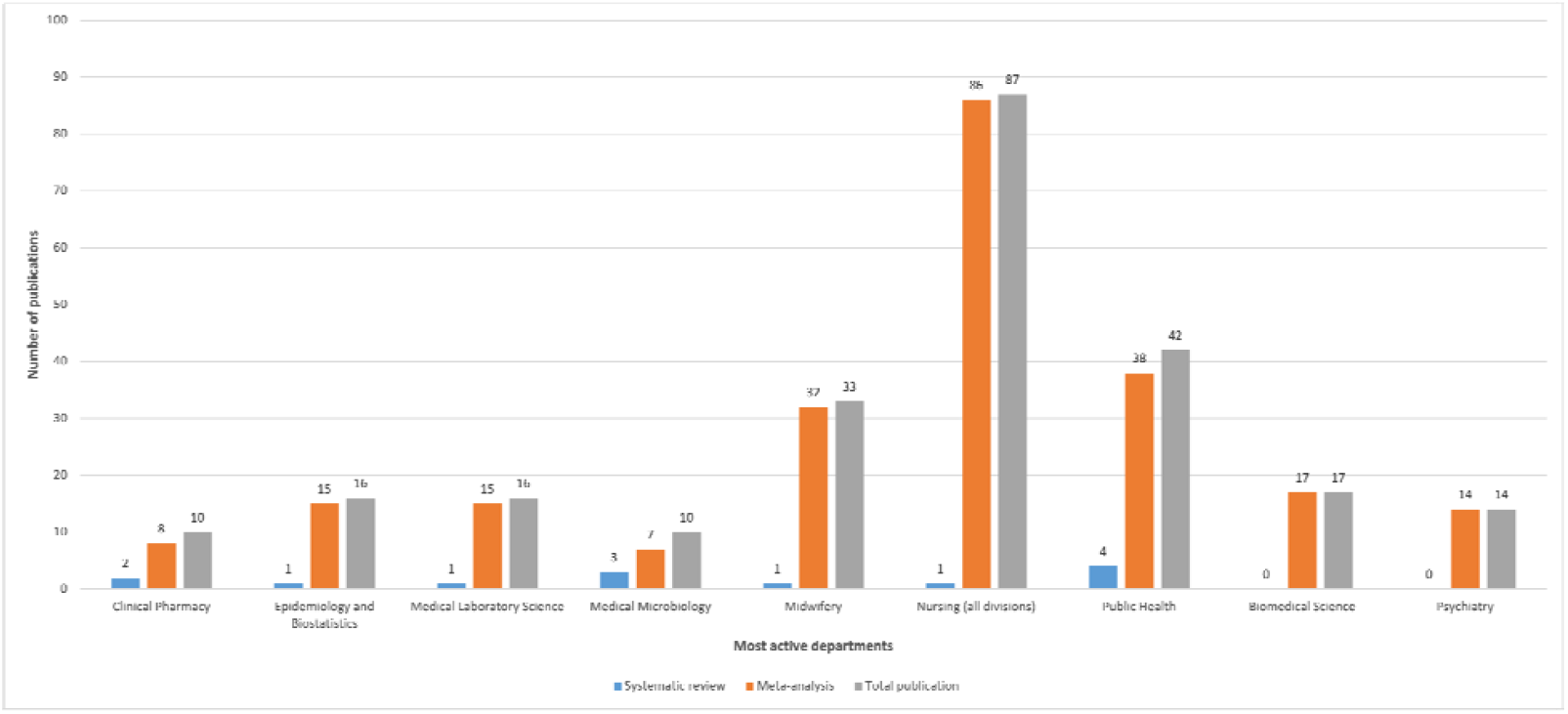
Most active departments publishing SR and MA.

**Fig 4.**
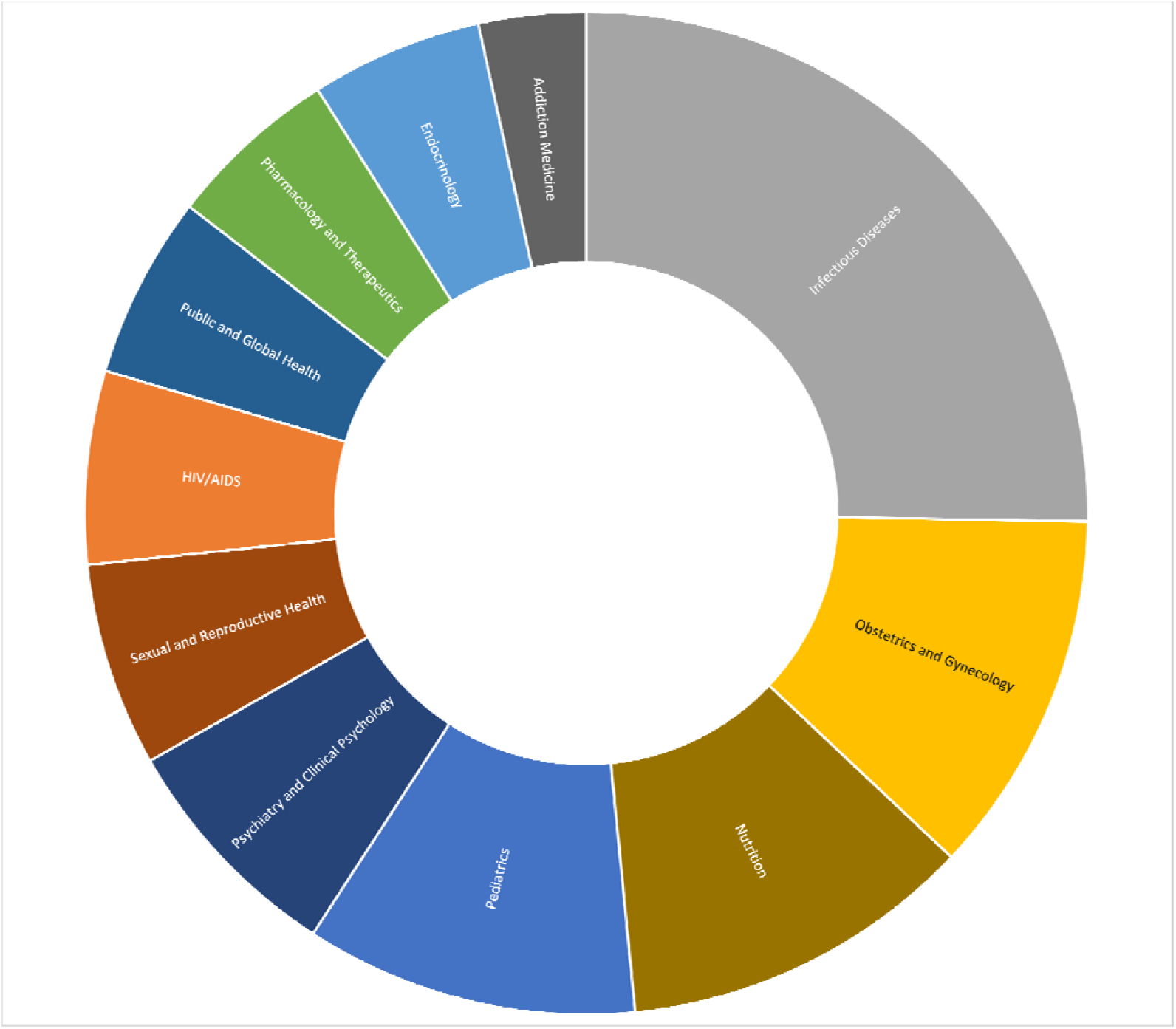
Most studied subject area.

MA articles were published in recent years (p <0.0001), registered their protocol (p <0.0001), authors were affiliated to multiple institutions (p = 0.007), published in high impact journals (p <0.01) and had high median number of references (p = 0.01) compared to the SR articles (Table 1). Conversely, SR had high median number of included studies (p = 0.003) and high median number of citation (p < 0.05) compared to MA (Table 1).

### Methodological and reporting quality

#### Protocol registration and searching strategy

As shown in Table 2 below, only 28.7% of SR and/or MA registered their protocol. Additionally, 10.3%, 27.5% and 24.4% did the searching without language or time restriction, did not use PICO or another standard searching framework and did not perform manual searching respectively. Moreover, 91.7% did not report the search interface, 63.6% did not report the search syntax and 77.9% did not report the date/year of the search coverage (Table 2).

**Table 2:**
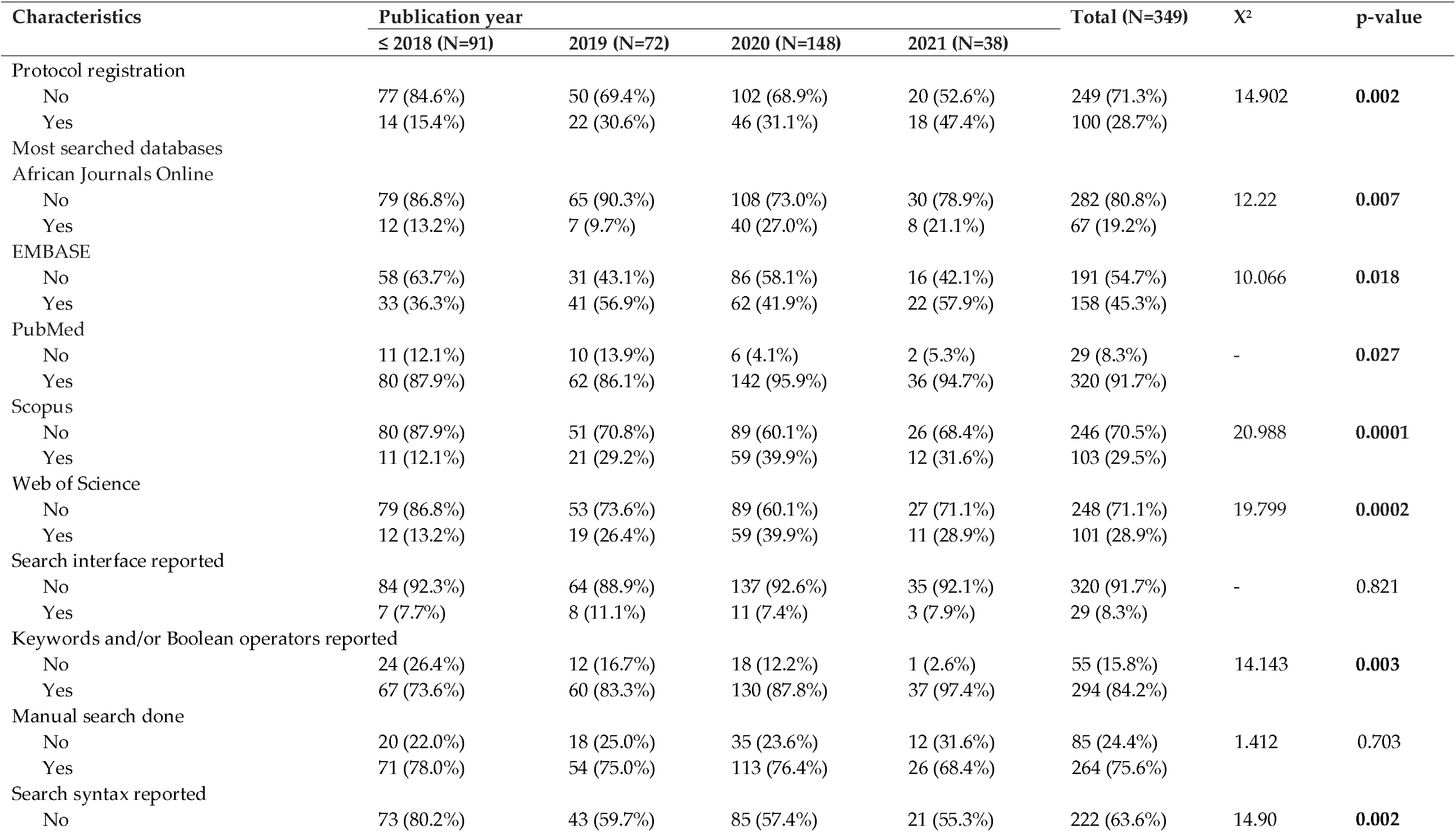
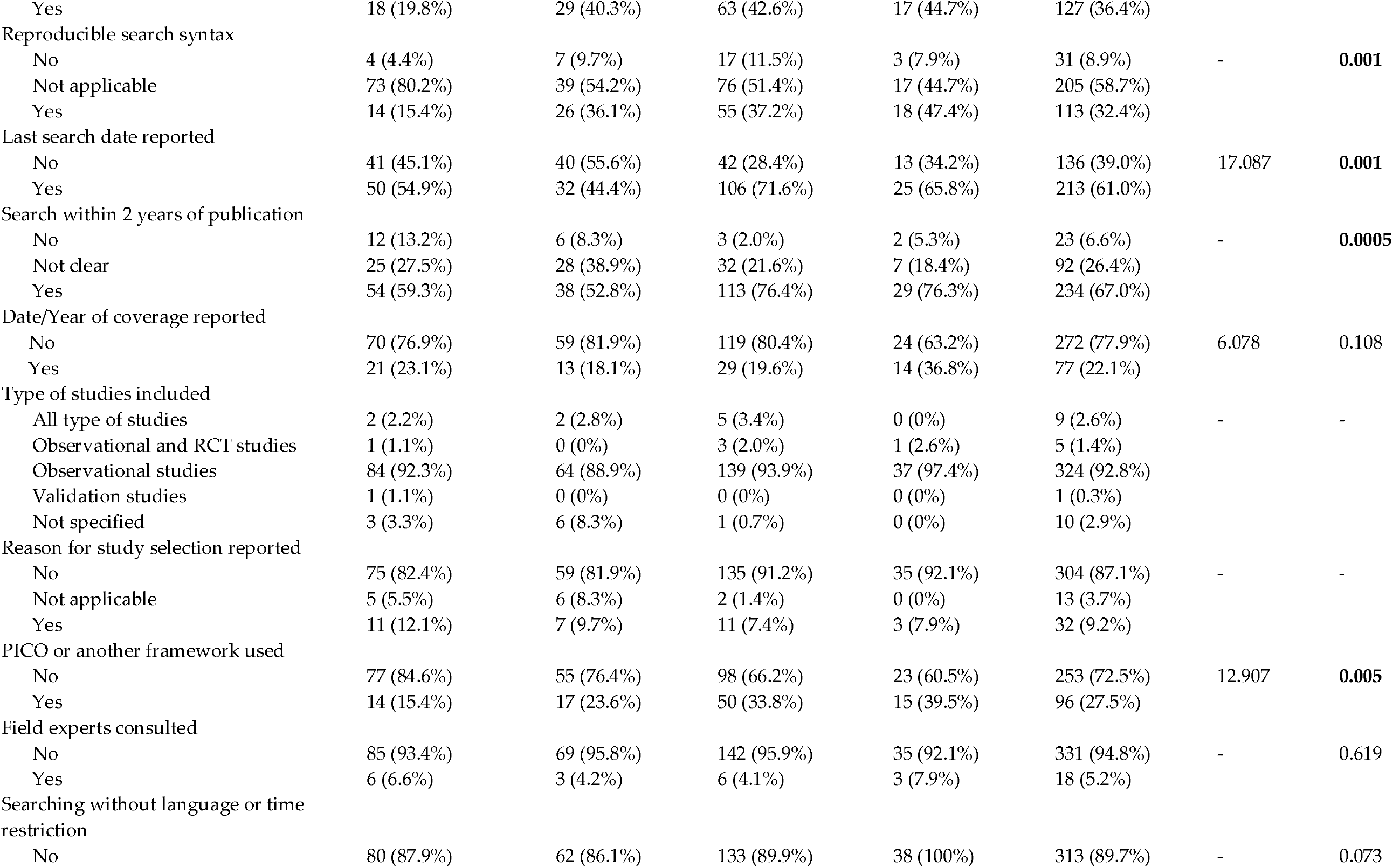
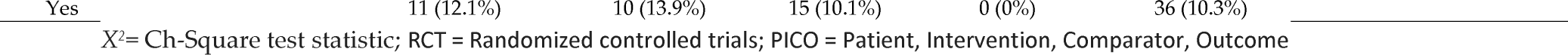
Protocol registration and searching strategy over time (N=349).

| Characteristics | Publication year |  |  |  | Total (N=349) | X<sup>2</sup> | p-value |
| --- | --- | --- | --- | --- | --- | --- | --- |
|  | ≤ 2018 (N=91) | 2019 (N=72) | 2020 (N=148) | 2021 (N=38) |  |  |  |
| Protocol registration |  |  |  |  |  |  |  |
| No | 77 (84.6%) | 50 (69.4%) | 102 (68.9%) | 20 (52.6%) | 249 (71.3%) | 14.902 | **0.002** |
| Yes | 14 (15.4%) | 22 (30.6%) | 46 (31.1%) | 18 (47.4%) | 100 (28.7%) |  |  |
| Most searched databases |  |  |  |  |  |  |  |
| African Journals Online |  |  |  |  |  |  |  |
| No | 79 (86.8%) | 65 (90.3%) | 108 (73.0%) | 30 (78.9%) | 282 (80.8%) | 12.22 | **0.007** |
| Yes | 12 (13.2%) | 7 (9.7%) | 40 (27.0%) | 8 (21.1%) | 67 (19.2%) |  |  |
| EMBASE |  |  |  |  |  |  |  |
| No | 58 (63.7%) | 31 (43.1%) | 86 (58.1%) | 16 (42.1%) | 191 (54.7%) | 10.066 | **0.018** |
| Yes | 33 (36.3%) | 41 (56.9%) | 62 (41.9%) | 22 (57.9%) | 158 (45.3%) |  |  |
| PubMed |  |  |  |  |  |  |  |
| No | 11 (12.1%) | 10 (13.9%) | 6 (4.1%) | 2 (5.3%) | 29 (8.3%) | - | **0.027** |
| Yes | 80 (87.9%) | 62 (86.1%) | 142 (95.9%) | 36 (94.7%) | 320 (91.7%) |  |  |
| Scopus |  |  |  |  |  |  |  |
| No | 80 (87.9%) | 51 (70.8%) | 89 (60.1%) | 26 (68.4%) | 246 (70.5%) | 20.988 | **0.0001** |
| Yes | 11 (12.1%) | 21 (29.2%) | 59 (39.9%) | 12 (31.6%) | 103 (29.5%) |  |  |
| Web of Science |  |  |  |  |  |  |  |
| No | 79 (86.8%) | 53 (73.6%) | 89 (60.1%) | 27 (71.1%) | 248 (71.1%) | 19.799 | **0.0002** |
| Yes | 12 (13.2%) | 19 (26.4%) | 59 (39.9%) | 11 (28.9%) | 101 (28.9%) |  |  |
| Search interface reported |  |  |  |  |  |  |  |
| No | 84 (92.3%) | 64 (88.9%) | 137 (92.6%) | 35 (92.1%) | 320 (91.7%) | - | 0.821 |
| Yes | 7 (7.7%) | 8 (11.1%) | 11 (7.4%) | 3 (7.9%) | 29 (8.3%) |  |  |
| Keywords and/or Boolean operators reported |  |  |  |  |  |  |  |
| No | 24 (26.4%) | 12 (16.7%) | 18 (12.2%) | 1 (2.6%) | 55 (15.8%) | 14.143 | **0.003** |
| Yes | 67 (73.6%) | 60 (83.3%) | 130 (87.8%) | 37 (97.4%) | 294 (84.2%) |  |  |
| Manual search done |  |  |  |  |  |  |  |
| No | 20 (22.0%) | 18 (25.0%) | 35 (23.6%) | 12 (31.6%) | 85 (24.4%) | 1.412 | 0.703 |
| Yes | 71 (78.0%) | 54 (75.0%) | 113 (76.4%) | 26 (68.4%) | 264 (75.6%) |  |  |
| Search syntax reported |  |  |  |  |  |  |  |
| No | 73 (80.2%) | 43 (59.7%) | 85 (57.4%) | 21 (55.3%) | 222 (63.6%) | 14.90 | **0.002** |
| Yes | 18 (19.8%) | 29 (40.3%) | 63 (42.6%) | 17 (44.7%) | 127 (36.4%) |  |  |
| Reproducible search syntax |  |  |  |  |  |  |  |
| No | 4 (4.4%) | 7 (9.7%) | 17 (11.5%) | 3 (7.9%) | 31 (8.9%) | - | **0.001** |
| Not applicable | 73 (80.2%) | 39 (54.2%) | 76 (51.4%) | 17 (44.7%) | 205 (58.7%) |  |  |
| Yes | 14 (15.4%) | 26 (36.1%) | 55 (37.2%) | 18 (47.4%) | 113 (32.4%) |  |  |
| Last search date reported |  |  |  |  |  |  |  |
| No | 41 (45.1%) | 40 (55.6%) | 42 (28.4%) | 13 (34.2%) | 136 (39.0%) | 17.087 | **0.001** |
| Yes | 50 (54.9%) | 32 (44.4%) | 106 (71.6%) | 25 (65.8%) | 213 (61.0%) |  |  |
| Search within 2 years of publication |  |  |  |  |  |  |  |
| No | 12 (13.2%) | 6 (8.3%) | 3 (2.0%) | 2 (5.3%) | 23 (6.6%) | - | **0.0005** |
| Not clear | 25 (27.5%) | 28 (38.9%) | 32 (21.6%) | 7 (18.4%) | 92 (26.4%) |  |  |
| Yes | 54 (59.3%) | 38 (52.8%) | 113 (76.4%) | 29 (76.3%) | 234 (67.0%) |  |  |
| Date/Year of coverage reported |  |  |  |  |  |  |  |
| No | 70 (76.9%) | 59 (81.9%) | 119 (80.4%) | 24 (63.2%) | 272 (77.9%) | 6.078 | 0.108 |
| Yes | 21 (23.1%) | 13 (18.1%) | 29 (19.6%) | 14 (36.8%) | 77 (22.1%) |  |  |
| Type of studies included |  |  |  |  |  |  |  |
| All type of studies | 2 (2.2%) | 2 (2.8%) | 5 (3.4%) | 0 (0%) | 9 (2.6%) | - | - |
| Observational and RCT studies | 1 (1.1%) | 0 (0%) | 3 (2.0%) | 1 (2.6%) | 5 (1.4%) |  |  |
| Observational studies | 84 (92.3%) | 64 (88.9%) | 139 (93.9%) | 37 (97.4%) | 324 (92.8%) |  |  |
| Validation studies | 1 (1.1%) | 0 (0%) | 0 (0%) | 0 (0%) | 1 (0.3%) |  |  |
| Not specified | 3 (3.3%) | 6 (8.3%) | 1 (0.7%) | 0 (0%) | 10 (2.9%) |  |  |
| Reason for study selection reported |  |  |  |  |  |  |  |
| No | 75 (82.4%) | 59 (81.9%) | 135 (91.2%) | 35 (92.1%) | 304 (87.1%) | - | - |
| Not applicable | 5 (5.5%) | 6 (8.3%) | 2 (1.4%) | 0 (0%) | 13 (3.7%) |  |  |
| Yes | 11 (12.1%) | 7 (9.7%) | 11 (7.4%) | 3 (7.9%) | 32 (9.2%) |  |  |
| PICO or another framework used |  |  |  |  |  |  |  |
| No | 77 (84.6%) | 55 (76.4%) | 98 (66.2%) | 23 (60.5%) | 253 (72.5%) | 12.907 | **0.005** |
| Yes | 14 (15.4%) | 17 (23.6%) | 50 (33.8%) | 15 (39.5%) | 96 (27.5%) |  |  |
| Field experts consulted |  |  |  |  |  |  |  |
| No | 85 (93.4%) | 69 (95.8%) | 142 (95.9%) | 35 (92.1%) | 331 (94.8%) | - | 0.619 |
| Yes | 6 (6.6%) | 3 (4.2%) | 6 (4.1%) | 3 (7.9%) | 18 (5.2%) |  |  |
| Searching without language or time restriction |  |  |  |  |  |  |  |
| No | 80 (87.9%) | 62 (86.1%) | 133 (89.9%) | 38 (100%) | 313 (89.7%) | - | 0.073 |

|  |  |  |  |  |  |
| --- | --- | --- | --- | --- | --- |
| Yes | 11 (12.1%) | 10 (13.9%) | 15 (10.1%) | 0 (0%) | 36 (10.3%) |

$X^2$ = Ch-Square test statistic; RCT = Randomized controlled trials; PICO = Patient, Intervention, Comparator, Outcome

Over time, we observed improvement in reporting of last search date (p = 0.001), search syntax (p = 0.002), and keywords and/or Boolean operators (p = 0.003) (Table 2). Also, we found that recent publications had a higher chance of having a registered protocol (p = 0.002), reproducible search syntax (p = 0.001), completed the search within two years of publication (p = 0.0005) and having used PICO or another standard framework for search (p = 0.005) (Table 2).

#### Screening and data extraction

The majority of SR and MA performed article screening (54.4%), data extraction (70.2%) and quality assessment (66.2%) in duplicate and improved over time (p < 0.0001). NOS (37.0%) was the most used quality assessment tool, followed by JBI (34.7%). Only 13.5% of the SR and MA used a standard (i.e., JBI) data extraction tool. Nearly all SR and MA (92.8%) included only observational studies, 87.1% did not mention the reason for study selection and 78.2% of the studies failed to contact or report contacting the corresponding authors for missing information or full-text. Moreover, 78.8% of the studies provided an adequate description of included studies and 91.7% reported the presence or absence of competing interest and/or funding for the SR or MA. However, most of the SRs and MA failed to report the list of excluded studies (86.8%) or funding for included primary studies (94.8%). Details has been presented in Table 3 below.

**Table 3:** Screening, data extraction and reporting over time (N=349).

| Characteristics | Publication year |  |  |  | Total (N=349) | X<sup>2</sup> | p-value |
| --- | --- | --- | --- | --- | --- | --- | --- |
|  | ≤ 2018 (N=91) | 2019 (N=72) | 2020 (N=148) | 2021 (N=38) |  |  |  |
| **Double screening for study selection** |  |  |  |  |  |  |  |
| No | 47 (51.6%) | 32 (44.4%) | 64 (43.2%) | 16 (42.1%) | 159 (45.6%) | 1.899 | 0.594 |
| Yes | 44 (48.4%) | 40 (55.6%) | 84 (56.8%) | 22 (57.9%) | 190 (54.4%) |  |  |
| **Double data extraction** |  |  |  |  |  |  |  |
| No | 46 (50.5%) | 24 (33.3%) | 25 (16.9%) | 9 (23.7%) | 104 (29.8%) | 31.626 | <0.0001 |
| Yes | 45 (49.5%) | 48 (66.7%) | 123 (83.1%) | 29 (76.3%) | 245 (70.2%) |  |  |
| **Data extraction tool** |  |  |  |  |  |  |  |
| Author(s) own (pretested) form | 49 (53.8%) | 40 (55.6%) | 100 (67.6%) | 28 (73.7%) | 217 (62.2%) | - | - |
| JBI | 8 (8.8%) | 8 (11.1%) | 24 (16.2%) | 7 (18.4%) | 47 (13.5%) |  |  |
| Not reported | 34 (37.4%) | 22 (30.6%) | 21 (14.2%) | 3 (7.9%) | 80 (22.9%) |  |  |
| **Contacting corresponding authors** |  |  |  |  |  |  |  |
| No | 71 (78.0%) | 62 (86.1%) | 111 (75.0%) | 29 (76.3%) | 273 (78.2%) | 3.616 | 0.306 |
| Yes | 20 (22.0%) | 10 (13.9%) | 37 (25.0%) | 9 (23.7%) | 76 (21.8%) |  |  |
| **Double quality assessment** |  |  |  |  |  |  |  |
| No | 47 (51.6%) | 24 (33.3%) | 33 (22.3%) | 14 (36.8%) | 118 (33.8%) | 21.868 | <0.0001 |
| Yes | 44 (48.4%) | 48 (66.7%) | 115 (77.7%) | 24 (63.2%) | 231 (66.2%) |  |  |
| **Mostly used quality assessment tools** |  |  |  |  |  |  |  |
| *JBI* |  |  |  |  |  |  |  |
| No | 47 (51.6%) | 33 (45.8%) | 79 (53.4%) | 21 (55.3%) | 180 (51.6%) | 2.654 | 0.448 |
| Yes | 22 (24.2%) | 23 (31.9%) | 60 (40.5%) | 16 (42.1%) | 121 (34.7%) |  |  |
| *NOS* |  |  |  |  |  |  |  |
| No | 48 (52.7%) | 28 (38.9%) | 79 (53.4%) | 17 (44.7%) | 172 (49.3%) | 7.414 | 0.060 |
| Yes | 21 (23.1%) | 28 (38.9%) | 60 (40.5%) | 20 (52.6%) | 129 (37.0%) |  |  |
| **List of excluded studies reported** |  |  |  |  |  |  |  |
| No | 73 (80.2%) | 63 (87.5%) | 134 (90.5%) | 33 (86.8%) | 303 (86.8%) | - | 0.162 |
| Yes | 17 (18.7%) | 9 (12.5%) | 13 (8.8%) | 5 (13.2%) | 44 (12.6%) |  |  |
| **Adequate description of included studies** |  |  |  |  |  |  |  |
| No | 28 (30.8%) | 18 (25.0%) | 23 (15.5%) | 5 (13.2%) | 74 (21.2%) | 9.918 | 0.019 |
| Yes | 63 (69.2%) | 54 (75.0%) | 125 (84.5%) | 33 (86.8%) | 275 (78.8%) |  |  |
| **Funding for included studies reported** |  |  |  |  |  |  |  |
| No | 90 (98.9%) | 68 (94.4%) | 136 (91.9%) | 37 (97.4%) | 331 (94.8%) | - | 0.090 |
| Yes | 1 (1.1%) | 4 (5.6%) | 12 (8.1%) | 1 (2.6%) | 18 (5.2%) |  |  |
| **Competing interest and/or funding reported** |  |  |  |  |  |  |  |
| No | 14 (15.4%) | 4 (5.6%) | 9 (6.1%) | 2 (5.3%) | 29 (8.3%) | - | 0.068 |
| Yes | 77 (84.6%) | 68 (94.4%) | 139 (93.9%) | 36 (94.7%) | 320 (91.7%) |  |  |

$X^2$ = Ch-Square test statistic; JBI = Joanna Briggs Institute; NOS = Newcastle-Ottawa Scale; PRISMA = Preferred Reporting Items for Systematic Reviews and Meta-Analyses

#### Statistical analyses methods

As shown in Table 4, most of the MA pooled prevalence estimates (78.8%) followed by odds ratio (47.6%) and estimates using random-effects model (78.8%). In 41.0% of the MA, DerSimonian-Laird method was used as between-study variance estimator and 84.5% used I^2^ statistic for heterogeneity assessment. Most of the MA used Egger’s test (72.8%) to assess publication bias and performed subgroup analysis (71.1%). STATA (66.2%) was the most used statistical software for meta-analyses.

**Table 4:**
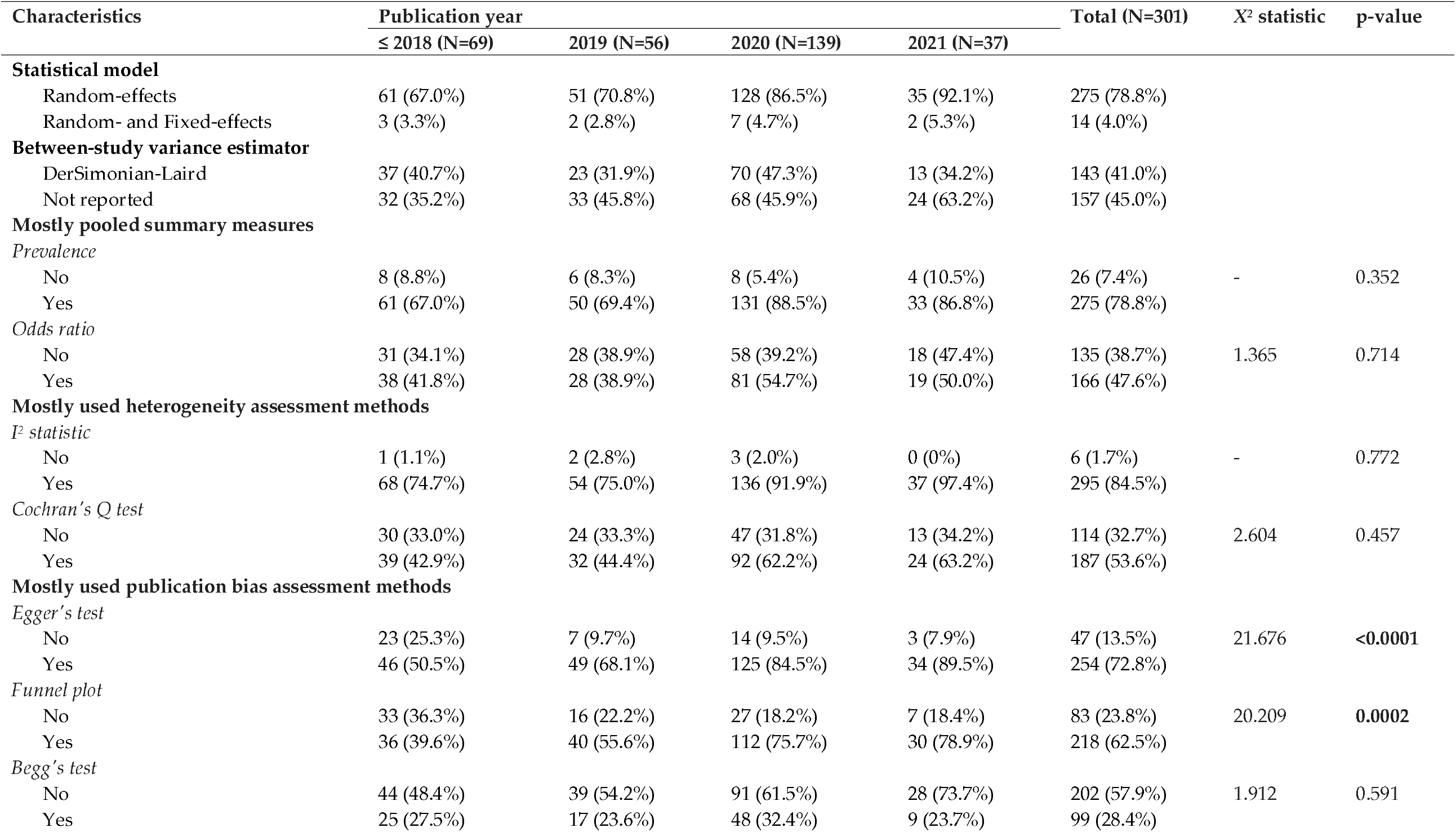
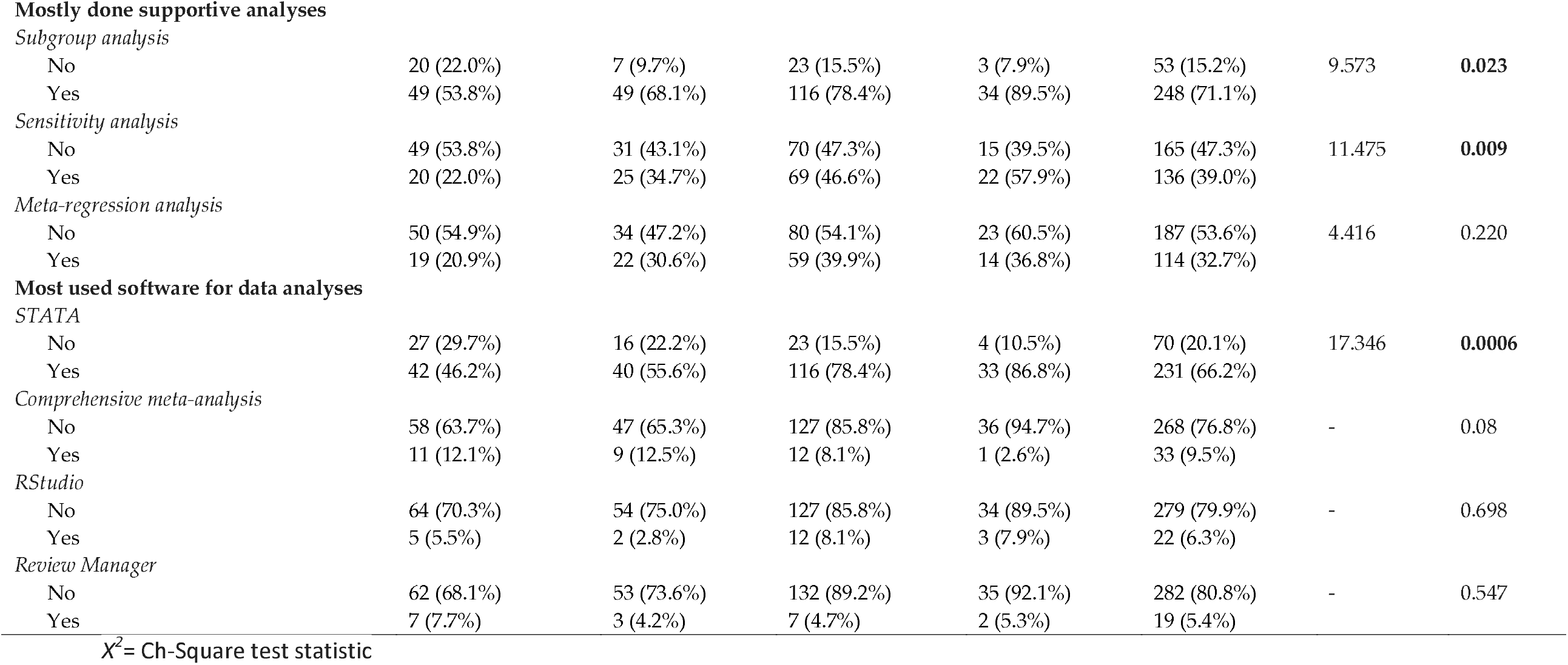
Statistical analyses methods used by MA (N=301)

| Characteristics | Publication year |  |  |  | Total (N=301) | X² statistic | p-value |
| --- | --- | --- | --- | --- | --- | --- | --- |
|  | ≤ 2018 (N=69) | 2019 (N=56) | 2020 (N=139) | 2021 (N=37) |  |  |  |
| **Statistical model** |  |  |  |  |  |  |  |
| Random-effects | 61 (67.0%) | 51 (70.8%) | 128 (86.5%) | 35 (92.1%) | 275 (78.8%) |  |  |
| Random- and Fixed-effects | 3 (3.3%) | 2 (2.8%) | 7 (4.7%) | 2 (5.3%) | 14 (4.0%) |  |  |
| **Between-study variance estimator** |  |  |  |  |  |  |  |
| DerSimonian-Laird | 37 (40.7%) | 23 (31.9%) | 70 (47.3%) | 13 (34.2%) | 143 (41.0%) |  |  |
| Not reported | 32 (35.2%) | 33 (45.8%) | 68 (45.9%) | 24 (63.2%) | 157 (45.0%) |  |  |
| **Mostly pooled summary measures** |  |  |  |  |  |  |  |
| *Prevalence* |  |  |  |  |  |  |  |
| No | 8 (8.8%) | 6 (8.3%) | 8 (5.4%) | 4 (10.5%) | 26 (7.4%) | - | 0.352 |
| Yes | 61 (67.0%) | 50 (69.4%) | 131 (88.5%) | 33 (86.8%) | 275 (78.8%) |  |  |
| *Odds ratio* |  |  |  |  |  |  |  |
| No | 31 (34.1%) | 28 (38.9%) | 58 (39.2%) | 18 (47.4%) | 135 (38.7%) | 1.365 | 0.714 |
| Yes | 38 (41.8%) | 28 (38.9%) | 81 (54.7%) | 19 (50.0%) | 166 (47.6%) |  |  |
| **Mostly used heterogeneity assessment methods** |  |  |  |  |  |  |  |
| *I² statistic* |  |  |  |  |  |  |  |
| No | 1 (1.1%) | 2 (2.8%) | 3 (2.0%) | 0 (0%) | 6 (1.7%) | - | 0.772 |
| Yes | 68 (74.7%) | 54 (75.0%) | 136 (91.9%) | 37 (97.4%) | 295 (84.5%) |  |  |
| *Cochran's Q test* |  |  |  |  |  |  |  |
| No | 30 (33.0%) | 24 (33.3%) | 47 (31.8%) | 13 (34.2%) | 114 (32.7%) | 2.604 | 0.457 |
| Yes | 39 (42.9%) | 32 (44.4%) | 92 (62.2%) | 24 (63.2%) | 187 (53.6%) |  |  |
| **Mostly used publication bias assessment methods** |  |  |  |  |  |  |  |
| *Egger's test* |  |  |  |  |  |  |  |
| No | 23 (25.3%) | 7 (9.7%) | 14 (9.5%) | 3 (7.9%) | 47 (13.5%) | 21.676 | <0.0001 |
| Yes | 46 (50.5%) | 49 (68.1%) | 125 (84.5%) | 34 (89.5%) | 254 (72.8%) |  |  |
| *Funnel plot* |  |  |  |  |  |  |  |
| No | 33 (36.3%) | 16 (22.2%) | 27 (18.2%) | 7 (18.4%) | 83 (23.8%) | 20.209 | 0.0002 |
| Yes | 36 (39.6%) | 40 (55.6%) | 112 (75.7%) | 30 (78.9%) | 218 (62.5%) |  |  |
| *Begg's test* |  |  |  |  |  |  |  |
| No | 44 (48.4%) | 39 (54.2%) | 91 (61.5%) | 28 (73.7%) | 202 (57.9%) | 1.912 | 0.591 |
| Yes | 25 (27.5%) | 17 (23.6%) | 48 (32.4%) | 9 (23.7%) | 99 (28.4%) |  |  |
| Subgroup analysis |  |  |  |  |  |  |  |
| No | 20 (22.0%) | 7 (9.7%) | 23 (15.5%) | 3 (7.9%) | 53 (15.2%) | 9.573 | **0.023** |
| Yes | 49 (53.8%) | 49 (68.1%) | 116 (78.4%) | 34 (89.5%) | 248 (71.1%) |  |  |
| Sensitivity analysis |  |  |  |  |  |  |  |
| No | 49 (53.8%) | 31 (43.1%) | 70 (47.3%) | 15 (39.5%) | 165 (47.3%) | 11.475 | **0.009** |
| Yes | 20 (22.0%) | 25 (34.7%) | 69 (46.6%) | 22 (57.9%) | 136 (39.0%) |  |  |
| Meta-regression analysis |  |  |  |  |  |  |  |
| No | 50 (54.9%) | 34 (47.2%) | 80 (54.1%) | 23 (60.5%) | 187 (53.6%) | 4.416 | 0.220 |
| Yes | 19 (20.9%) | 22 (30.6%) | 59 (39.9%) | 14 (36.8%) | 114 (32.7%) |  |  |
| STATA |  |  |  |  |  |  |  |
| No | 27 (29.7%) | 16 (22.2%) | 23 (15.5%) | 4 (10.5%) | 70 (20.1%) | 17.346 | **0.0006** |
| Yes | 42 (46.2%) | 40 (55.6%) | 116 (78.4%) | 33 (86.8%) | 231 (66.2%) |  |  |
| Comprehensive meta-analysis |  |  |  |  |  |  |  |
| No | 58 (63.7%) | 47 (65.3%) | 127 (85.8%) | 36 (94.7%) | 268 (76.8%) | - | 0.08 |
| Yes | 11 (12.1%) | 9 (12.5%) | 12 (8.1%) | 1 (2.6%) | 33 (9.5%) |  |  |
| RStudio |  |  |  |  |  |  |  |
| No | 64 (70.3%) | 54 (75.0%) | 127 (85.8%) | 34 (89.5%) | 279 (79.9%) | - | 0.698 |
| Yes | 5 (5.5%) | 2 (2.8%) | 12 (8.1%) | 3 (7.9%) | 22 (6.3%) |  |  |
| Review Manager |  |  |  |  |  |  |  |
| No | 62 (68.1%) | 53 (73.6%) | 132 (89.2%) | 35 (92.1%) | 282 (80.8%) | - | 0.547 |
| Yes | 7 (7.7%) | 3 (4.2%) | 7 (4.7%) | 2 (5.3%) | 19 (5.4%) |  |  |

$X^2$ = Ch-Square test statistic

Across the publication years, we observed an increased trend in using Egger’s test (p <0.0001) and funnel plot (p-value 0.0002) for assessing publication bias. Similarly, performing subgroup analysis (p = 0.023) and sensitivity analysis (p = 0.009) increased over time (Table 4).

## Discussion

This study described the epidemiological trends and methodological and reporting quality of SR and MA in medicine and health sciences in Ethiopia. We found that the publication rate has been increased over time, especially during the last three years. We also observed a promising methodological and reporting quality improvements over time, even though there is a long way to go.

Since 2018, the number of SR and MA publications in Ethiopia has been dramatically increased with 148 SR and MA published in 2020. This increase is in line with the global trend in several fields. For example, a recent study found that about 80 SR were globally published per day in 2019 – a 20-fold increase from the number 20 years ago.^3^ The increase in the publication of SR and MA in Ethiopia maybe because SR and MA can be completed in short time period compared to primary studies. Also, the development of free citation management and statistical analysis software with freely accessible meta-analysis commands may have likely contributed to its increase, which saves substantial amount of time. Moreover, the increasing recognition of SR and MA by the academic institutions, scientific community, and healthcare practitioners as a more reliable source of evidence can motivate authors. The increase in number of higher institutions, and MSc and PhD graduates in recent years further support our findings. Coupled with the fact that SR and MA do not require acquisition of new primary data—which are challenged by limited resources, the need for ethical approval, and feasibility of conducting multicenter studies—SR and MA are better alternatives for many researchers in low resource settings like Ethiopia.

The implication of the increased SR and MA publication is double-edged. On the one hand, it is useful to produce up-to-date knowledge for evidence-based decision-making to improve health and safety and save costs. On the other hand, there is also a growing concern that the mass production of SR and MA might lead to unnecessary, misleading, and conflicting when not properly conducted.^21-24^ Most healthcare decision-making tools were based on evidence from developed countries without due consideration of the cultural and socioeconomic differences, and local context.^25^ Moreover, a high number of SR might lead to research waste, as the publication of overlapping SR and MA is becoming evident.^26, 27^ For example, we found 28 duplicates on various topics: two on mother-to-child transmission of HIV^28, 29^, two on HIV/AIDS treatment failure^30, 31^, three on nursing process^32-34^, three on antenatal depression^35-37^, three on postnatal depression^38-40^, three on breastfeeding^41-43^, three on immunization coverage^44-46^, three on low birth weight^47-49^, two on tuberculosis treatment non-adherence^50, 51^, two on maternal-near miss^52, 53^ and two on antiepleptive medication non-adherence.^54, 55^ To minimize research waste, authors must register their protocol, thoroughly search protocol registration cites and communicate each other whenever possible. This can also increase collaboration and save energy and time.

Despite the increased trend of SR and MA publication in Ethiopia, most were poorly conducted and reported when evaluated based on the international standard criteria. The common methodological drawbacks in most SR and MA were not having a registered protocol and not reporting the search interface, search syntax, last search date, and list of excluded studies. This might be related to the quality of publishing journals. In this study, we observed that more than 50% of SR and MA were published in the lower half of the journal groups – Q3 and Q4 or unranked journals, which they may not be strict about the quality of SR and MA, and publish all submitted manuscripts to increase their journal impact. Furthermore, authors may publish SR and MA just out of interest without adequate knowledge and skill or consulting methodologists and statisticians. Our findings are consistent with the global reports, where inconsistencies in methodological and reporting quality of SR and MA is commonly reported.^5^ Ensuring methodological and reporting quality is a long-standing challenge for researchers.^6^ This implies that authors, peer reviewers and journal editors should work towards publishing high standard SR and MA. It is advisable for authors to look back and invite co-authors or other friends to read their manuscript instead of rushing to publish low quality SR and MA, and institutions must evaluate the quality of evidence before using these articles for academic promotion. Additionally, journal editors might consider asking authors to include their SR protocol along with the manuscript submission, and check for their appropriateness before peer review invitation. Moreover, to ensure quality, funding agencies would evaluate experience and expertise in SR and MA of grant applicants and proposal reviewers and may request documentation of SR and MA training or publication.

On the other hand, we observed a promising trend of improvement in some aspects of qualities in methodology and reporting in SR and MA in Ethiopia. Over time, there was an improved registration of protocols, searching more databases and transparently reporting search strategy. This agrees with previous studies that reported improvement in methodological quality over time.^4, 5^ This may be due to increase authors knowledge and skills to conduct SR and MA. In our study, we observed that at least four out of ten SR and MA published in Q1 (top 25%) and Q2 (50 - 75%) journals, which we believe their quality is better than SR and MA published in low rank or unranked journals. Furthermore, the improvement in reporting can be attributed to the development of methodological and reporting guidelines, such as the PRISMA statement^56^ and the AMSTAR checklist.^17^

Our study’s main strength is that we included a large number of SR and MA published to date, which enabling us to observe publication trends and current status of quality. In addition, we gathered detailed background- and quality-related data, which provides authors the opportunity to identify main quality-related setbacks and start prioritizing quality over quantity. Our study also has some limitations. First, we did not include unpublished SRs and MA, such as preprints, in our analysis. Given the fact that the peer reviewer and editorial feedback would affect the quality of articles, we felt that preprints may not be sufficient for assessing methodological ore reporting quality. Second, misclassification for subject categories is possible and there is a huge inconsistency in choosing the subject area by authors in medrxiv platform. In addition, some outcomes may have more than one category, for example, H-pylori infection can be an infectious disease and gastroenterology. Third, only 10% of the data is validated by independent reviewer.

## Conclusions

For the first time, our study provides robust characterization of epidemiological trends and methodological and reporting quality of SR and MA published to date in Ethiopia. Although the publication rate of SR and MA has been increased over time, it was not balanced with the increase in methodological and reporting quality. Most authors did not register their protocol and did not report search interface, search syntax, last search date, and list of excluded studies. Overall, we observed a promising improvement in some aspects of methodological and reporting quality of SR and MA. Shared responsibility between authors, peer reviewers, editors, journals and publishers in optimizing quality of the outputs through adhering to the accepted methodological quality and using appropriate reporting templates.

## Data Availability

All data produced in the present study are available upon reasonable request to the authors

https://osf.io/q5dw2/

---

**Supplementary Table 1:**
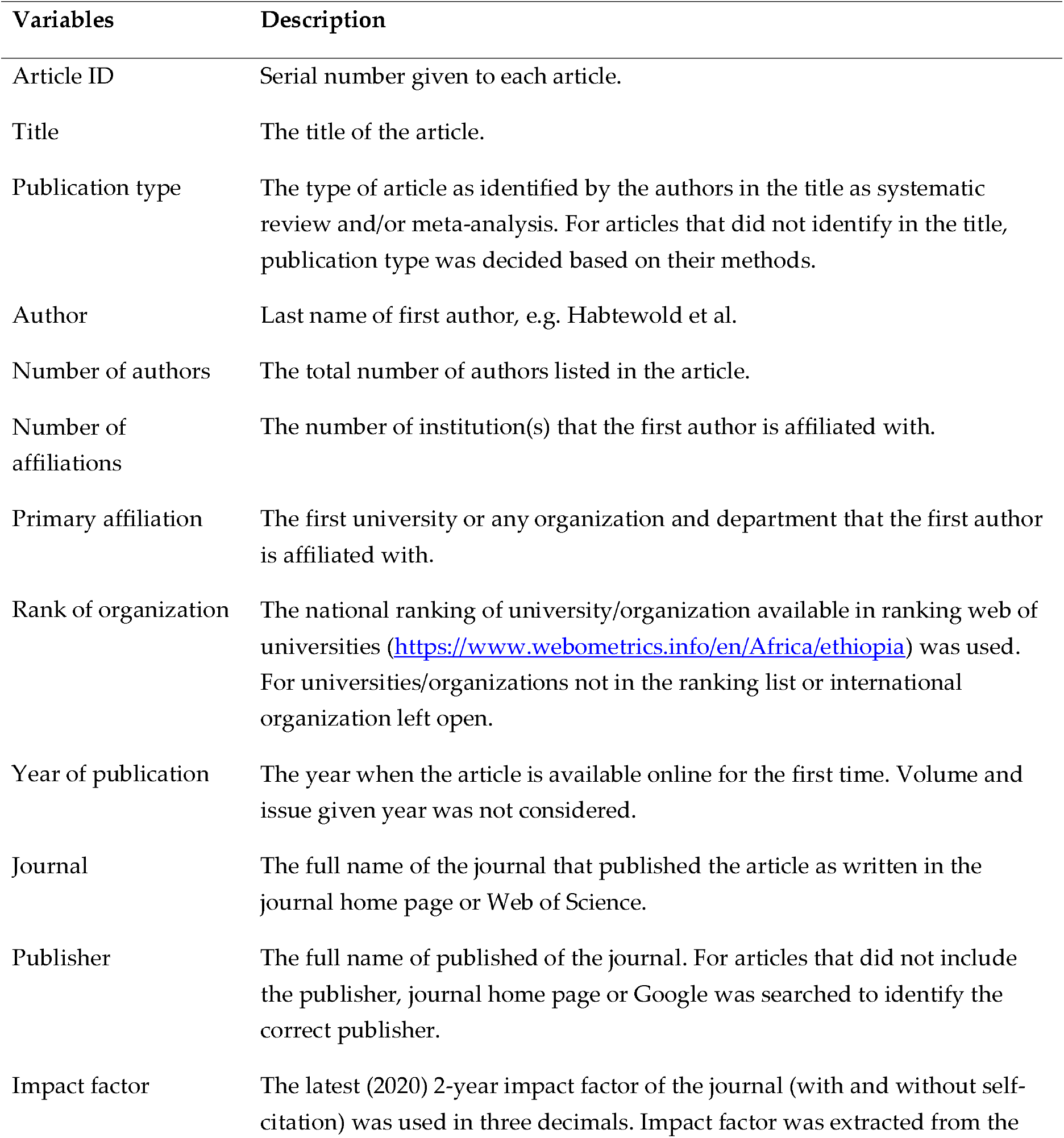
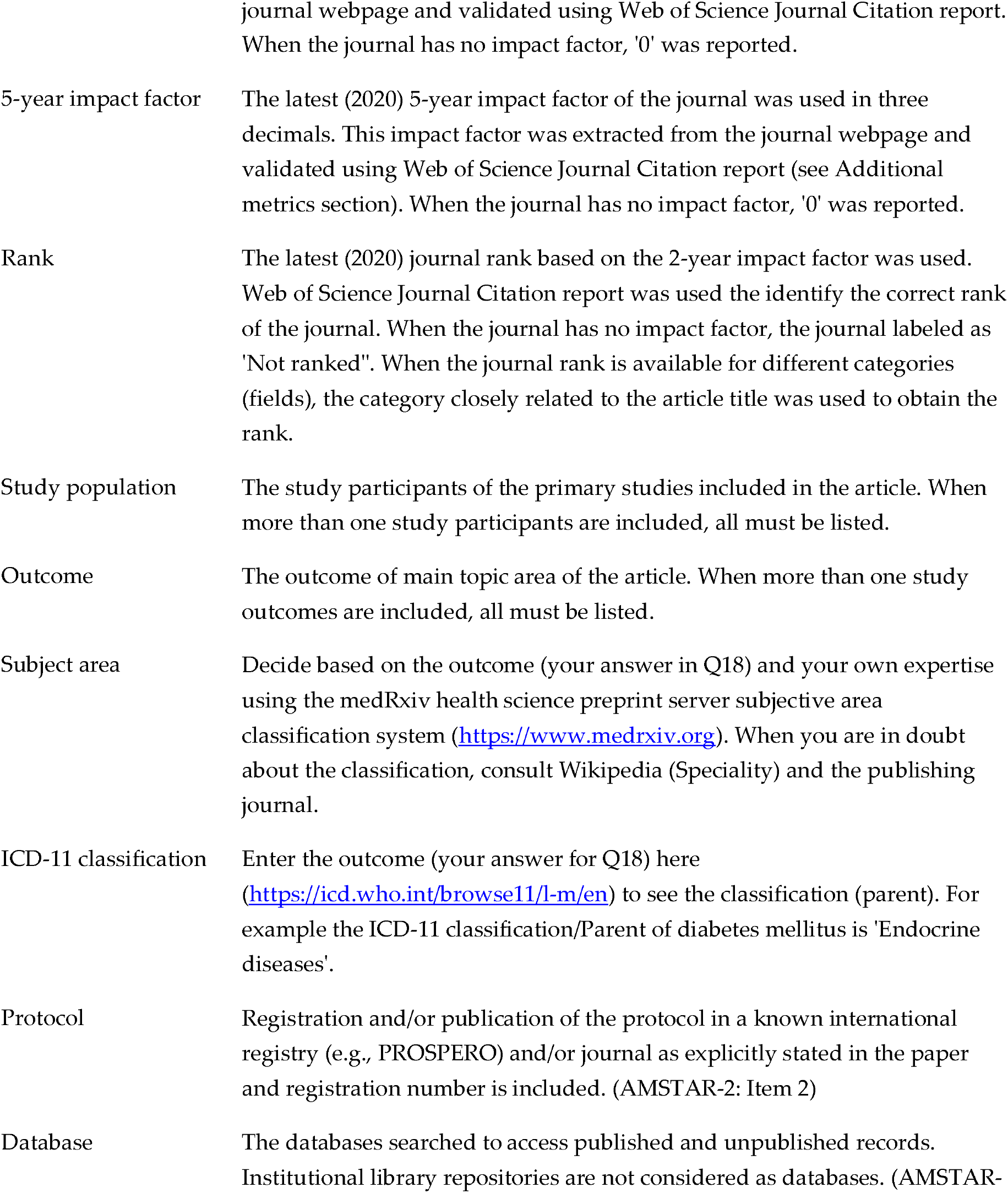
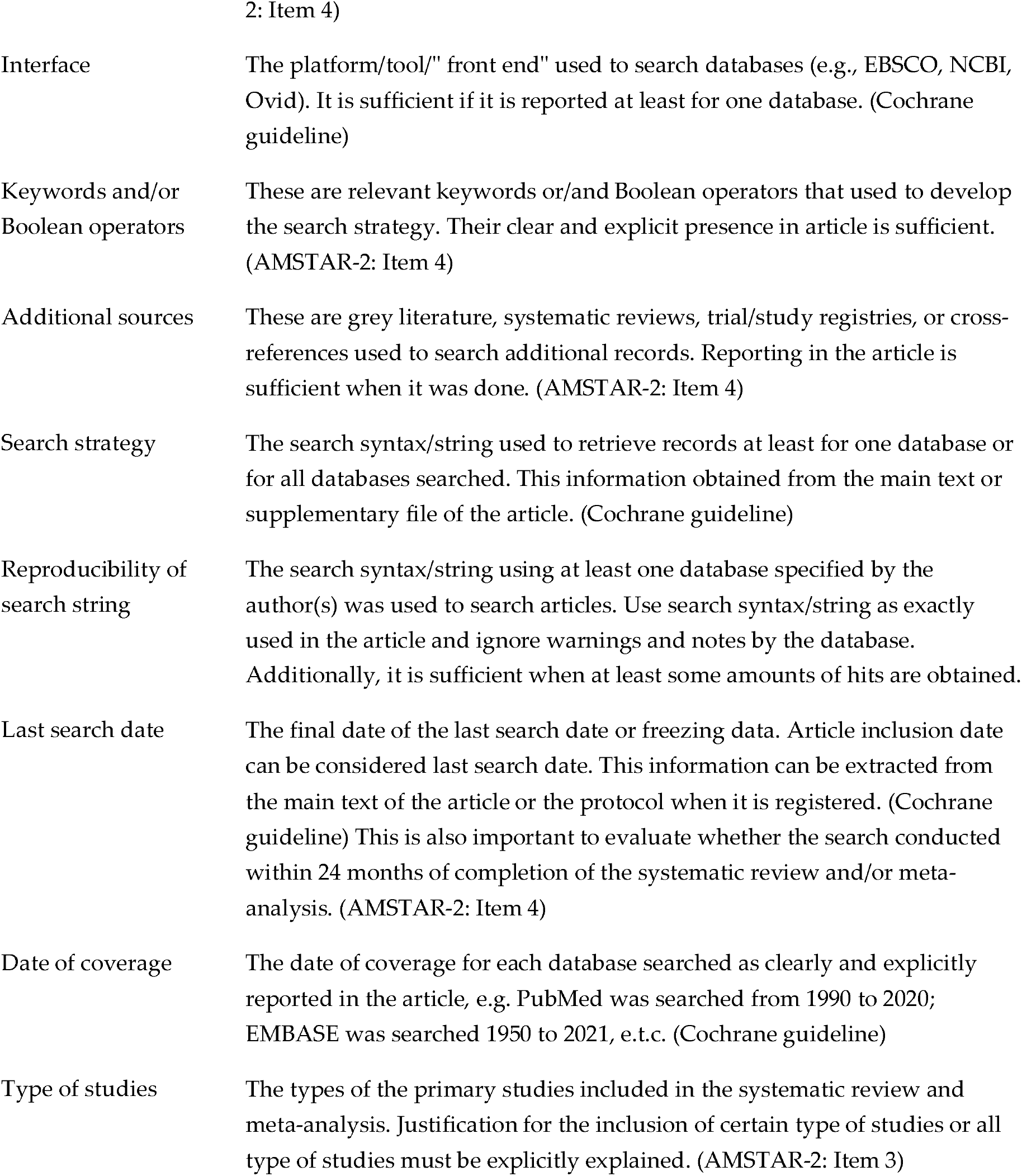
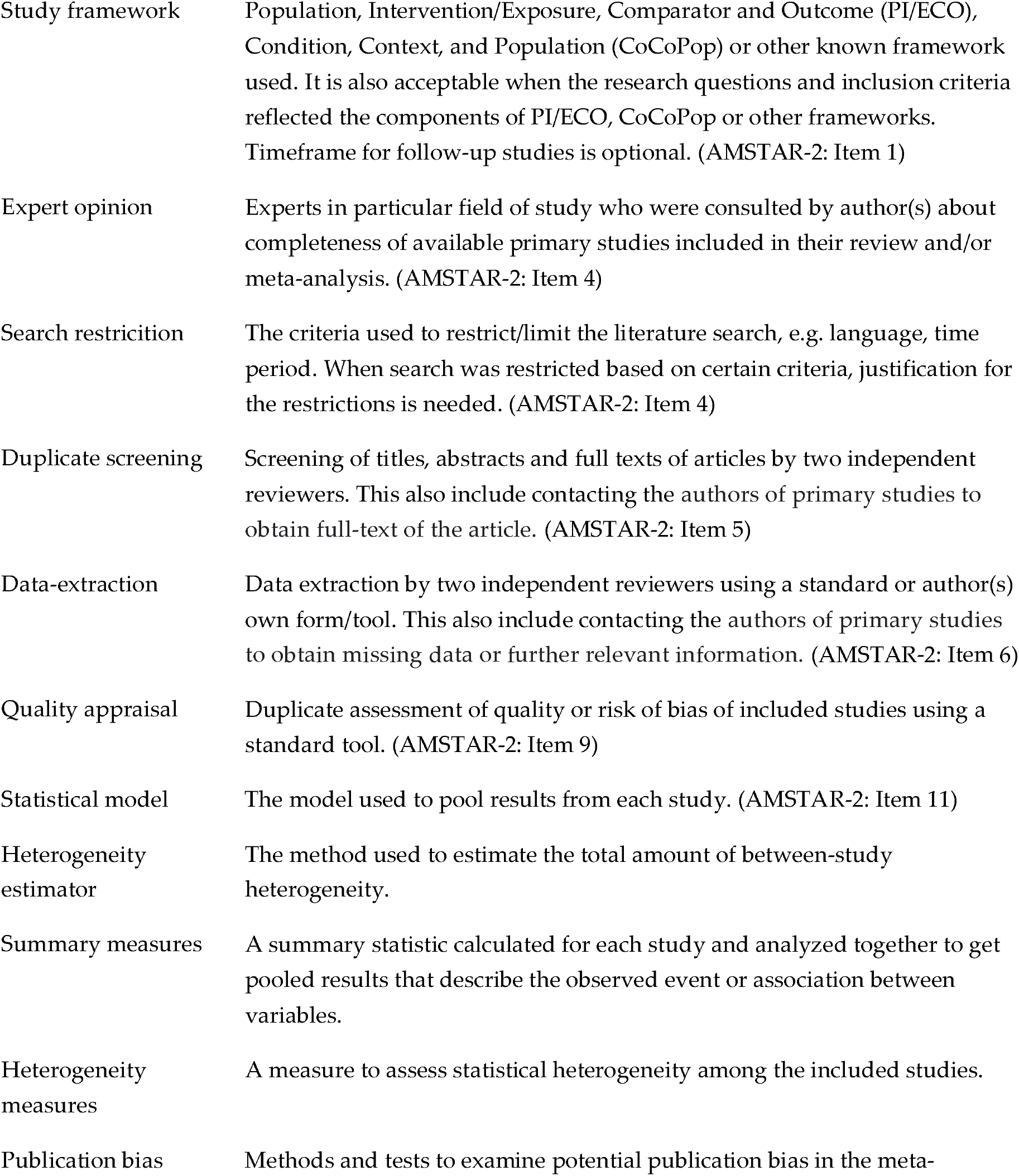
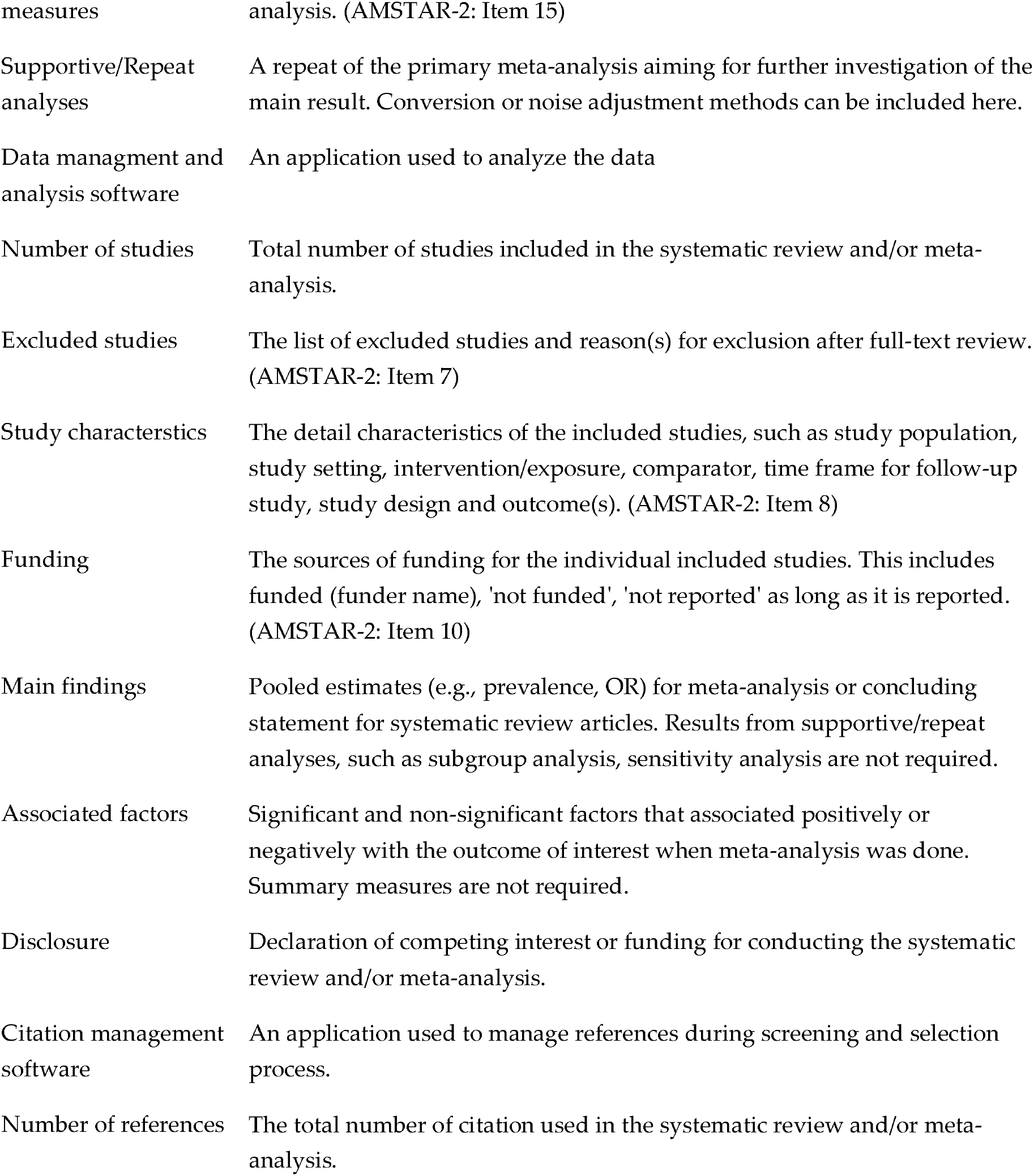
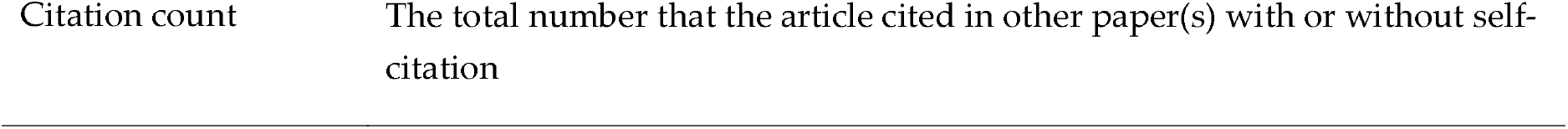
List and description of variables extracted from systematic reviews and meta-analyses.

## References

1. Fusar-Poli P, Radua J. Ten simple rules for conducting umbrella reviews. Evid Based Ment Health Aug 2018;21(3):95–100.

2. Hossain MM. Umbrella review as an emerging approach of evidence synthesis in health sciences: a bibliometric analysis. Available at SSRN 3551055 2020.

3. Hoffmann F, Allers K, Rombey T, Helbach J, Hoffmann A, Mathes T, Pieper D. Nearly 80 systematic reviews were published each day: Observational study on trends in epidemiology and reporting over the years 2000-2019. Journal of Clinical Epidemiology 2021/10/01/ 2021;138:1–11.

4. Moher D, Tetzlaff J, Tricco AC, Sampson M, Altman DG. Epidemiology and reporting characteristics of systematic reviews. PLoS Med Mar 27 2007;4(3):e78.

5. Page MJ, Shamseer L, Altman DG, et al. Epidemiology and Reporting Characteristics of Systematic Reviews of Biomedical Research: A Cross-Sectional Study. PLOS Medicine 2016;13(5):e1002028.

6. Yoshii A, Plaut DA, McGraw KA, Anderson MJ, Wellik KE. Analysis of the reporting of search strategies in Cochrane systematic reviews. J Med Libr Assoc 2009;97(1):21–29.

7. Tian J, Zhang J, Ge L, Yang K, Song F. The methodological and reporting quality of systematic reviews from China and the USA are similar. Journal of Clinical Epidemiology 2017/05/01/ 2017;85:50–58.

8. Bougioukas KI, Vounzoulaki E, Mantsiou CD, Papanastasiou GD, Savvides ED, Ntzani EE, Haidich A-B. Global mapping of overviews of systematic reviews in healthcare published between 2000 and 2020: a bibliometric analysis. Journal of Clinical Epidemiology 2021/09/01/ 2021;137:58–72.

9. Habtewold TD, Alemu SM, Mohammed SH, et al. Biomedical and public health reviews and meta-analyses in Ethiopia had poor methodological quality: overview of evidence from 1970 to 2018. Journal of Clinical Epidemiology 2019/05/01/ 2019;109:90–98.

10. Mohammed SH, Habtewold TD, Arero AG, Esmaillzadeh A. The state of child nutrition in Ethiopia: an umbrella review of systematic review and meta-analysis reports. BMC Pediatr Aug 26 2020;20(1):404.

11. Bayih WA, Birhane BM, Belay DM, et al. The state of birth asphyxia in Ethiopia: An umbrella review of systematic review and meta-analysis reports, 2020. Heliyon Oct 2021;7(10):e08128.

12. Habtewold TD, Sharew NT, Endalamaw A, et al. Bibliometric study of preclinical, clinical, and public health systematic reviews and meta-analyses in Ethiopia: systematically mapping publication outputs, authors’ collaboration networks, trending research topics, and influential articles. medRxiv 2022:2022.2002.2024.22271416.

13. Higgins JP, Thomas J, Chandler J, Cumpston M, Li T, Page MJ, Welch VA. Cochrane handbook for systematic reviews of interventions: John Wiley & Sons; 2019.

14. Page MJ, McKenzie JE, Bossuyt PM, et al. The PRISMA 2020 statement: An updated guideline for reporting systematic reviews. J Clin Epidemiol Jun 2021;134:178–189.

15. Hupe M. EndNote X9. Journal of Electronic Resources in Medical Libraries 2019;16(3-4):117–119.

16. Covidence. Covidence systematic review software, Veritas Health Innovation, Melbourne, Australia.

17. Shea BJ, Reeves BC, Wells G, et al. AMSTAR 2: a critical appraisal tool for systematic reviews that include randomised or non-randomised studies of healthcare interventions, or both. Bmj Sep 21 2017;358:j4008.

18. Aromataris E, Fernandez RS, Godfrey C, Holly C, Khalil H, Tungpunkom P. Methodology for JBI umbrella reviews. 2014.

19. Faggion CM, Jr., Wu YC, Tu YK, Wasiak J. Quality of search strategies reported in systematic reviews published in stereotactic radiosurgery. Br J Radiol Jun 2016;89(1062):20150878.

20. Corbyons K, Han J, Neuberger MM, Dahm P. Methodological Quality of Systematic Reviews Published in the Urological Literature from 1998 to 2012. J Urol Nov 2015;194(5):1374–1379.

21. Ioannidis JP. The Mass Production of Redundant, Misleading, and Conflicted Systematic Reviews and Meta-analyses. Milbank Q Sep 2016;94(3):485–514.

22. Page MJ, Moher D. Mass Production of Systematic Reviews and Meta-analyses: An Exercise in Mega-silliness? Milbank Q Sep 2016;94(3):515–519.

23. Grainger MJ, Bolam FC, Stewart GB, Nilsen EB. Evidence synthesis for tackling research waste. Nature Ecology & Evolution 2020/04/01 2020;4(4):495–497.

24. Nakagawa S, Dunn AG, Lagisz M, et al. A new ecosystem for evidence synthesis. Nature Ecology & Evolution 2020/04/01 2020;4(4):498–501.

25. Pearson A, Jordan Z. Evidence-based healthcare in developing countries. Int J Evid Based Healthc Jun 2010;8(2):97–100.

26. Siontis KC, Hernandez-Boussard T, Ioannidis JPA. Overlapping meta-analyses on the same topic: survey of published studies. BMJ : British Medical Journal 2013;347:f4501.

27. Naudet F, Schuit E, Ioannidis JPA. Overlapping network meta-analyses on the same topic: survey of published studies. Int J Epidemiol Dec 1 2017;46(6):1999–2008.

28. Kassa GM. Mother-to-child transmission of HIV infection and its associated factors in Ethiopia: a systematic review and meta-analysis. BMC Infect Dis May 10 2018;18(1):216.

29. Endalamaw A, Demsie A, Eshetie S, Habtewold TD. A systematic review and meta-analysis of vertical transmission route of HIV in Ethiopia. BMC Infect Dis Jun 22 2018;18(1):283.

30. Endalamaw A, Mekonnen M, Geremew D, Yehualashet FA, Tesera H, Habtewold TD. HIV/AIDS treatment failure and associated factors in Ethiopia: meta-analysis. BMC Public Health Jan 20 2020;20(1):82.

31. Assemie MA, Alene M, Ketema DB, Mulatu S. Treatment failure and associated factors among first line patients on highly active antiretroviral therapy in Ethiopia: a systematic review and meta-analysis. Glob Health Res Policy 2019;4:32.

32. Gebeyehu Yazew K, Azagew AW, Yohanes YB. Determinants of the nursing process implementation in Ethiopia: A systematic review and meta-analysis, 2019. International Journal of Africa Nursing Sciences 2020/01/01/ 2020;13:100219.

33. Shiferaw WS, Akalu TY, Wubetu AD, Aynalem YA. Implementation of Nursing Process and Its Association with Working Environment and Knowledge in Ethiopia: A Systematic Review and Meta-Analysis. Nurs Res Pract 2020;2020:6504893.

34. Bayih WA, Ayalew MY, Belay DM, et al. The implementation of nursing process during patient care in Ethiopia: A systematic review and meta-analysis. Heliyon May 2021;7(5):e06933.

35. Zegeye A, Alebel A, Gebrie A, Tesfaye B, Belay YA, Adane F, Abie W. Prevalence and determinants of antenatal depression among pregnant women in Ethiopia: a systematic review and meta-analysis. BMC Pregnancy Childbirth Nov 29 2018;18(1):462.

36. Ayano G, Tesfaw G, Shumet S. Prevalence and determinants of antenatal depression in Ethiopia: A systematic review and meta-analysis. PLoS One 2019;14(2):e0211764.

37. Getinet W, Amare T, Boru B, Shumet S, Worku W, Azale T. Prevalence and Risk Factors for Antenatal Depression in Ethiopia: Systematic Review. Depress Res Treat 2018;2018:3649269.

38. Duko B, Wolde D, Alemayehu Y. The epidemiology of postnatal depression in Ethiopia: a systematic review and meta-analysis. Reprod Health Nov 19 2020;17(1):180.

39. Desta M, Memiah P, Kassie B, Ketema DB, Amha H, Getaneh T, Sintayehu M. Postpartum depression and its association with intimate partner violence and inadequate social support in Ethiopia: a systematic review and meta-analysis. J Affect Disord Jan 15 2021;279:737–748.

40. Necho M, Abadisharew M, Getachew Y. A Systematic Review and Meta-analysis of Depression in Postpartum Women in a Low-income Country; Ethiopia, 2020. The Open Public Health Journal 2020;13(1).

41. Habtewold TD, Mohammed SH, Endalamaw A, et al. Breast and complementary feeding in Ethiopia: new national evidence from systematic review and meta-analyses of studies in the past 10 years. Eur J Nutr Oct 2019;58(7):2565–2595.

42. Alebel A, Tesma C, Temesgen B, Ferede A, Kibret GD. Exclusive breastfeeding practice in Ethiopia and its association with antenatal care and institutional delivery: a systematic review and meta-analysis. Int Breastfeed J 2018;13:31.

43. Alebel A, Dejenu G, Mullu G, Abebe N, Gualu T, Eshetie S. Timely initiation of breastfeeding and its association with birth place in Ethiopia: a systematic review and meta-analysis. Int Breastfeed J 2017;12:44.

44. Biset G, Woday A, Mihret S, Tsihay M. Full immunization coverage and associated factors among children age 12-23 months in Ethiopia: systematic review and meta-analysis of observational studies. Hum Vaccin Immunother Jul 3 2021;17(7):2326–2335.

45. Eshete A, Shewasinad S, Hailemeskel S. Immunization coverage and its determinant factors among children aged 12-231months in Ethiopia: a systematic review, and Meta-analysis of cross-sectional studies. BMC Pediatr Jun 8 2020;20(1):283.

46. Nour TY, Farah AM, Ali OM, Abate KH. Immunization coverage in Ethiopia among 12-231month old children: systematic review and meta-analysis. BMC Public Health Jul 20 2020;20(1):1134.

47. Endalamaw A, Engeda EH, Ekubagewargies DT, Belay GM, Tefera MA. Low birth weight and its associated factors in Ethiopia: a systematic review and meta-analysis. Ital J Pediatr Nov 26 2018;44(1):141.

48. Dasa TT, Kassie TW, Roba AA, Kelel HU. Prevalence and Determinants of Low Birth Weight in Ethiopia: A Systematic Review and Meta-Analysis. African Journal of Health Sciences 2020;33(2):49–64.

49. Katiso NA, Kassa GM, Fekadu GA, Kidanemariam Berhe A, Muche AA. Prevalence and Determinants of Low Birth Weight in Ethiopia: A Systematic Review and Meta-Analysis. Advances in Public Health 2020/09/15 2020;2020:7589483.

50. Zegeye A, Dessie G, Wagnew F, Gebrie A, Islam SMS, Tesfaye B, Kiross D. Prevalence and determinants of anti-tuberculosis treatment non-adherence in Ethiopia: A systematic review and meta-analysis. PLoS One 2019;14(1):e0210422.

51. Tola HH, Holakouie-Naieni K, Tesfaye E, Mansournia MA, Yaseri M. Prevalence of tuberculosis treatment non-adherence in Ethiopia: a systematic review and meta-analysis. Int J Tuberc Lung Dis Jun 1 2019;23(6):741–749.

52. Turi E, Fekadu G, Taye B, et al. The impact of antenatal care on maternal near-miss events in Ethiopia: A systematic review and meta-analysis. International Journal of Africa Nursing Sciences 2020/01/01/ 2020;13:100246.

53. Mengist B, Desta M, Tura AK, Habtewold TD, Abajobir A. Maternal near miss in Ethiopia: Protective role of antenatal care and disparity in socioeconomic inequities: A systematic review and meta-analysis. International Journal of Africa Nursing Sciences 2021/01/01/ 2021;15:100332.

54. Amha H, Memiah P, Getnet A, et al. Antiseizure medication nonadherence and its associated factors among Epileptic patients in Ethiopia, a systematic review and meta-analysis. Seizure Oct 2021;91:462–475.

55. Belayneh Z, Mekuriaw B. A systematic review and meta-analysis of anti-epileptic medication non-adherence among people with epilepsy in Ethiopia. Arch Public Health 2020;78:23.

56. Moher D, Liberati A, Tetzlaff J, Altman DG. Preferred reporting items for systematic reviews and meta-analyses: the PRISMA statement. PLoS Med Jul 21 2009;6(7):e1000097.

